# The Impact of Non-Price In-premise Marketing on Food and Beverage Purchasing and Consumer Behaviour: A Systematic Review

**DOI:** 10.1101/2021.09.13.21258115

**Authors:** R. Whitehead, S. Greci, H Thomson, G. Armour, K. Angus, L. Martin

## Abstract

In-premise marketing is commonly used to promote foods that are high in fat, sugar or salt. In order to inform development of public policy in this area, this systematic review sought to determine the quantity and quality of English-language evidence which examines the role and impact of in-premise advertising (e.g., signage, posters) and positional promotions (e.g., checkout displays) on consumer behaviour and diet-related outcomes in retail, out-of-home (i.e., cafés, restaurants, takeaways) and online purchasing environments. Sixty-two studies met inclusion criteria, of which 69% (n=42) were identified as being methodologically weak. The best-available evidence constitutes findings from four methodologically strong studies, and ten moderate studies which are not confounded by additional promotions such as price or availability. These studies predominantly found evidence that in-premise marketing is likely to be successful in influencing consumer behaviour towards targeted items, across retail and out-of-home settings. These findings provide a basis for authorities to consider acting to restrict in-premise marketing of unhealthy foods and encouraging the in-premise marketing of healthier products. This review identified gaps in the evidence available on non-sales outcomes, and on online purchase environments. These gaps, and identified methodological limitations of the extant evidence remain to be addressed by future research.

## Introduction

The World Health Organisation’s 2004 global strategy on diet, physical activity and health^1^ identified marketing within the food environment as one of the main determinants of food choice and dietary habits. This strategy recommended that national governments and the private sector act to implement responsible marketing practices of foods that are high in fat, sugar or salt (HFSS), in order to reduce the burden of disease attributable to obesity. In 2010 this was followed by specific recommendations to reduce children’s exposure to marketing of less healthy foods.^2^ However, despite these calls, obesity and marketing of unhealthy foods remain major challenges in high income countries.^3,4^

In-premise marketing seeks to elicit impulsive, preferential consumer responses towards targeted products. This is achieved by appealing to automatic information processing and disrupting more deliberative cognitive processing mechanisms. This can involve increasing visual salience, increasing perceived value, creating feelings of urgency and shifting consumers’ reference frames.^5^ There is also evidence that the pervasiveness of food marketing in this context may act to distort perceptions of normal food purchasing behaviour, by increasing the social acceptability of targeted items and normalising purchases of larger quantities of food and drink.^6^ As these marketing approaches are widely utilised for unhealthy foods^7–9^, they can frequently act to disrupt individuals’ capacity in their own long-term self-interests. As such, there is a strong argument to consider regulatory action, to make it easier for individuals to make intentional, rather than reflexive food choices.

Marketing strategies typically encompass a combination of four elements relating to the product, its price, promotion and placement.^10^ The role of price in particular is well understood, with changes in price being reliably and quantifiably associated with the elasticity of demand.^11^ Less clear, however, is the role and impact of in-premise advertising (e.g., in-premise signage and posters, shelf-edge signage, promotion of value, verbal prompts) and positional promotions (e.g., checkout displays, end-of-aisle displays, special island displays). These aspects of the marketing mix are often used to promote HFSS foods, with evidence suggesting they are particularly salient to young people.^12^

Review-level literature on the impact of in-premise marketing is sparse and has tended to focus on price and availability.^13–15^ Whilst one recent systematic review^13^ found moderate evidence that prominent positioning within a retail environment is associated with dietary outcomes, no prior systematic reviews have specifically investigated the impact of both in-premise advertising and positional promotion across the entirety of the commercial food environment, including retail, out-of-home (i.e., cafés, restaurants, takeaways) and online settings. The out-of-home environment is of particular interest in this context given evidence that up to 25% of caloric intake is consumed out of home^16^ and findings from store audits^17^ that promotions in this sector predominantly target discretionary (i.e., foods providing little or no nutritional benefit) and HFSS foods. Out-of-home marketing of unhealthy foods also stands to maintain socioeconomic health inequalities as deprived areas tend to have a greater density of, and easier accessibility to takeaway and fast-food restaurants.^18^ The role of the online food environment is of increasing importance given secular trends in shopping habits,^19^ which have been accelerated by the COVID-19 pandemic.^20–23^ This review, therefore, aims to address the following questions across retail, out-of-home and online shopping environments, in order to inform the development of public policy:

1. What research evidence is available on non-price in-premise marketing (defined for this review as: positional promotions, in-store advertising and equivalent online promotions) of food and beverages?
2. What is the quality of the available evidence which has assessed the impact of non-price in-premise marketing of food and beverages on consumer behaviour or diet-related outcomes?
3. What is the impact of non-price in-premise marketing of food and beverages on consumer behaviour (including: consumer purchasing, consumer preferences, intention to purchase, consumer attention) or diet-related outcomes.

## Methods

The protocol for this review was registered with the International Prospective Register for Systematic Reviews (PROSPERO): CRD42020169720. The review methods and findings are reported according to PRISMA^24^ and synthesis without meta-analysis (SWiM)^25^ guidelines.

Studies were sought and included in this review where they examined an exposure or intervention which altered in-premise positioning or advertising of any food or non-alcoholic drink product in a retail, out-of-home or online setting. For the purposes of this review, in-premise positional promotions included: checkout displays, end-of-aisle displays, front of store displays and special island/bin displays. In-premise advertising promotions include shelf edge signage, in-premise posters, branded chillers, promotion of value, stand-alone offers and verbal prompts. Equivalent promotions in an online environment were considered, including pop-up prompts, front page placement and banner advertisements. Additional inclusion and exclusion criteria are detailed in Table 1.

Nine electronic databases were searched (Business Source Complete; Cochrane Central Register of Controlled Trials; Cochrane Database of Systematic Reviews; EconLit; Emerald Insight; PsycINFO; SAGE Business Cases; Web of Science Core Collection; World Advertising Research Centre). All databases were searched from inception to 01 May 2020. The search strategy was developed by experienced information scientists (GA & KA) and included terms relating to “marketing”, “advertising”, “promotion”, “retail”, “out of home”, “online”, “food” and “drink” were used to identify studies that described the association between in-premise marketing and outcomes such as sales, consumer preference, customer attention, dietary quality and/or body mass index (BMI). The full search strategies used for each database are presented as supporting information (S1).

Using Covidence ©,^26^ all titles and abstracts were independently screened by two authors (LM, RW or SG) against inclusion criteria. Disagreement was resolved either via discussion or by a casting vote by a third reviewer. If it was unclear from the abstract alone whether a study was eligible for inclusion, the full text was reviewed. The full text of each potentially relevant study identified through title and abstract screening was then independently reviewed by two authors (LM, RW or SG), again with disagreement resolved via discussion or a casting vote. Exclusions at full-text review were hierarchically applied and are reported in the flow chart (Figure 1). If, following full-text review it was unclear whether a study met inclusion criteria, the corresponding author was emailed to ask for clarification. This was necessary in three cases, with two authors providing the information within four weeks. Studies were excluded as “unobtainable” where the requested information was not provided. Bibliographies of 37 existing systematic reviews were also screened for additional studies. This process identified one additional study for inclusion.

Details regarding included study characteristics (study setting, participant details, food/ drink type, exposures, outcomes) were independently extracted by two reviewers (LM & RW) and recorded using Covidence ©. Only results that were deemed to be relevant to the research question were extracted during the data synthesis process. Reviewers compared the data extracted for consistency and discussed to reach consensus.

Quality appraisal was completed independently in duplicate (LM & RW) to determine the risk of bias using the Effective Public Health Practice Project (EPHPP) tool.^27^ This tool assesses eight bias domains including participant selection, study design, handling of confounders, blinding, data collection, drop-outs, intervention fidelity and statistical analyses. Studies were graded as either weak (two or more weak domains), moderate (one weak domain) or strong (no weak domains). Discrepancies in overall score between the two reviewers were discussed, with discussion of discrepant domain and individual item assessments as necessary until an overall quality score was agreed.

Synthesis was primarily conducted by grouping eligible studies by each permutation of setting (retail, out-of-home and online) and outcome category (sales, consumer preference, attention, dietary quality and health status). Where studies provide results pertinent to more than one setting, outcome, or intervention, they are considered independently in separate syntheses. A separate grouping was conducted on the basis of the directness of intervention evidence. This involved grouping studies which examine the impact of changes to promotional activity designed to actively *reduce* consumer responsiveness to less healthy foods. These reflect the most direct assessment of any policies that may be put in place to attempt to reduce population consumption of HFSS and discretionary foods. These more direct studies are prioritised in relation to the remaining studies which reflect a relatively indirect assessment of potential policy responses in this context. Within all groupings, where available, studies identified as being at lower risk of bias were also prioritised when drawing conclusions. Furthermore, it became clear that some studies allowed the examination of in-premise advertising and/or positional promotions in isolation, whereas others were confounded by concurrent use of promotions that are outside of the scope of this review (e.g., price and availability). The presence of these ‘confounding promotions’ means we can be less certain that promotions of interest are responsible for any observed association with outcomes, therefore, in addition to prioritisation on the basis of directness and methodological quality, studies were also grouped and additionally prioritised where they allow examination of in-scope promotions in isolation (either through the study design or statistical controlling). Prioritised studies are grouped and presented earlier, and in greater detail in syntheses and are given greater weight when reaching overarching conclusions.

A meta-analysis of standardized effect size estimates was not possible due to the heterogeneity of the study designs, interventions, statistical reporting and outcomes. As such, a vote counting method based on direction of effect (as recommended in the Cochrane handbook)^28,29^ was used to summarise the effect direction and statistical significance of findings for each outcome and setting permutation on a per-study basis (i.e., if a study included multiple results for similar outcomes in the same setting, the results were aggregated). Per study, all relevant findings were aggregated and labelled as an increase or decrease where 70% or more were in the same direction, or inconsistent where this threshold was not met. Similarly, findings were labelled as statistically significant if 70% or more of the study’s relevant findings were significant at p<.05, non-significant if 70% or more were non-significant at p>.05 and inconsistent in other cases. Where multiple results of interest are reported, only those allowing the most direct examination of in-scope promotional strategies were selected (i.e., as close as possible to a comparison between two groups or conditions where the sole difference is the presence or absence of in-premise marketing). Extracted data on effect direction are reported narratively and in effect direction plots (Supporting information S2-6). Effect direction but not statistical significance was used to reach conclusions across studies, in line with Cochrane guidance.^29^

## Results

### Characteristics of included studies

Search results and the screening process are presented in Figure 1. Sixty-two studies met inclusion criteria,^30–91^ of which 33 present research conducted in a retail environment, 24 in an out-of-home environment and six in an online shopping environment. One study^43^ examined the impact of an intervention based in both retail and out-of-home contexts and as such is considered in syntheses across each of these settings below. Additional characteristics of the included studies are presented in detail in supporting information (S2-6), and are briefly summarised below.

Half of the included studies (31/62) present research conducted in the United States, 9 studies (15%) in the United Kingdom, 7 (11%) in the Netherlands. Eleven studies (18%) collected data from elsewhere in Europe, including one cross-national study.^47^ The remaining research was conducted in Australia (n=2, 3%) and Canada (n=2, 3%).

The included studies were inconsistent in their provision of information on sample size, meaning formal comparison on this basis (in terms of either number of premises, transactions or participants) was not possible. Any pertinent information available in this regard is presented in supporting information (S2-6). Information on consumers’ demographic characteristics was also inconsistently reported, with very few identified studies formally assessing differential impacts of in-premise marketing across demographic groups such as age, sex or socioeconomic status.

The methodological quality of the included literature was predominantly poor, with over two thirds of the included studies (43 of 62) being assessed as weak according to the EPHPP tool.^27^ Fifteen studies (24%) were assessed as being of moderate quality, with the remaining four (6%) obtaining a strong rating. Twenty three (37%) of the included studies used a randomised, controlled design. Fifteen studies (24%) used a non-randomised controlled design, 15 (24%) an uncontrolled pre-post design and nine (15%) an observational design.

Included studies predominantly (n=48, 77%) assessed the association between in-premise marketing and sales outcomes, with these data chiefly being recorded electronically at point of sale (n=30). Other means of recording sales outcomes included participant self-report (n=7), researcher observation (n=6), collection of sales receipts (n=2), analysis of take-home panel data (n=2) and storeowner report (n=1). Ten studies (16%) measured consumer preferences for promoted items, including three recording purchase intentions. Five studies (8%) recorded outcomes relating to consumption, via self-report (n=3), researcher observation (n=1) and weighing of plate waste (n=1). Three studies (5%) estimated participants’ macronutrient intake using self-reported food frequency questionnaires. One study (2%) assessed the association between in-premise marketing and BMI.^59^ One study (2%) recorded the association between in-premise marketing and customers’ attention.^57^ Five studies collected data in more than one outcome category (4 with two types of outcome^32,37,51,59^ one with three^43^), these studies are considered separately in each relevant synthesis below.

Twenty-three of the included studies (37%) evaluated the impact of positional promotions, with 44 (71%) evaluating types of in-premise advertising. Eleven (18%) of the included studies evaluated the impact of both of these types of promotion employed simultaneously. Six studies (10%) evaluated the impact of advertisements in an online shopping environment. The most commonly observed positional promotions include checkout placement (n=15), end-of-aisle displays (n=6) and special island displays (n=6). A wide range of in-premise advertising was observed including posters or signage (n=24), shelf labelling (n=10), text and formatting of menus (n=9) and table tents (n=3). In-premise marketing largely targeted healthier foods (n=48, 77%), with fewer included studies examining promotions targeting discretionary (n=12, 19%) or wider HFSS foods (n=8, 13%).

Eight studies were identified that explored interventions designed to reduce (rather than promote) the prominence of a product.^44,46,54,60,71,74,86,91^ Four of these achieved this via the removal of a product from prominent locations (checkouts), whilst keeping it available elsewhere within the premises.^44,54,71,74^ A further four studies used a displacement approach, whereby adding a product to a prominent position aims to reduce the salience of another within the display (for example aiming to reduce sales of confectionery at a checkout by introducing fruit to this location).^46,60,86,91^ Six studies analysed the impact of both promotional interventions seeking to increase the sales of some products and reduce the prominence of other products.^44,54,60,71,86,91^ Findings from these studies are considered separately in syntheses for both active promotion and restrictive or displacement interventions below.

Over half of the included studies (n=36, 58%) allowed examination of the impact of promotions included within the scope of this review in isolation from confounding promotions. In the remaining 26 studies (42%), the unique impact of in-scope promotions is impossible to examine in isolation from confounding promotions, the most commonly observed of which were increased product availability (12 studies), placement of a product within a shelf or aisle (7 studies), price (5 studies) and taste testing (5 studies).

### Association with outcomes

#### Restrictive interventions

Four studies^44,54,71,74^ investigated physical removal of products from prominent positions in a retail setting (whilst maintaining their availability elsewhere in the premises), providing a direct examination of the potential impact of restricting positional promotions. These studies are detailed in supporting information (S2) and overall all found that removal of products from checkout displays was associated with a reduction in sales^54,71,74^ or an increase in reported dietary quality,^44^ with this association statistically significant in two studies.

The best available evidence of the impact of restrictive interventions comes from one methodologically strong study using an observational, interrupted time series design.^74^ This study used a nationally representative panel of around 30,000 UK households to assess, in isolation of confounding interventions, the impact of changes in national supermarket policy. The introduction of policies to remove discretionary foods from checkout displays was found to be associated with a significant ∼17% reduction in purchases of these items.

#### Displacement interventions

Four studies assessed the impact of in-premise marketing on sales of items that themselves are not the target of promotion (i.e., a displacement effect, for example the effect of active promotion of fruit at a checkout on sales of confectionary sold in this location) in retail^46,60^ and out-of-home^86,91^ premises (supporting information, S3). Overall, three of these^46,60,91^ found that prominent positioning or in-premise advertising of healthier products was associated with a reduction in sales of less healthy items (despite no change in the positioning or availability of the latter category of products). One study^86^ found this approach to be associated with a non-significant increase in sales of confectionery. There is little evidence of these associations being statistically significant, with only one study^91^ finding inconsistent evidence that an observed decrease in sales of confectionary and croissants (following promotion of fruit and bread products) was statistically significant.

The best available evidence of displacement interventions’ impact comes from one moderate quality study, without confounding promotions.^46^ This study, using a non-randomised controlled design, found that supermarkets with checkout displays promoting fruit, vegetables, water, nut and cereal bars observed a non-significant reduction in confectionary sales, relative to control stores.

### Retail promotions

Overall, evidence of in-premise marketing’s positive influence on consumer behaviour (e.g., increased sales, consumption or preferences of targeted items) was seen in 26 of 35 observed outcomes (across 33 studies, see supporting information, S4) in a retail setting, with this association significant in 9 studies. One study^58^ found evidence that a retail promotion had a non-significant negative influence on consumer behaviour (reduced sales), and eight found inconsistent evidence with respect to effect direction.

#### Retail promotions: Sales outcomes

Twenty-seven studies examined the effect of in-premise marketing on sales in a retail environment (supporting information, S4). Overall, 21 (78%) of these found that in-premise marketing was associated with an increase in sales (7 of which were statistically significant, 3 provided inconsistent evidence of statistical significance). Five studies found inconsistent evidence on the direction of this association, and one weak study^58^ found evidence that in-premise advertising was associated with a non-significant reduction in sales.

Two methodologically strong studies^70,73^ were included which assess in-premise promotions’ impact on sales in a retail setting, both allowing examination of positional promotions^73^ or in-premise advertising^70^ in isolation from confounding promotions. One of these^70^ found that for two out of three products (olive oil and coffee) supermarket shelf labels were associated with a significant increase in sales, but for a third (flour) shelf labelling was associated with a non-significant decrease. The second study^73^ found that end-of-aisle displays in supermarkets were associated with a significant increase in the sales of carbonated drinks, coffee and tea.

Three studies of moderate quality^68,79,90^ allowed examination of positional promotions in isolation from confounding promotions. These each found that positional promotions were significantly associated with an increase in sales of promoted items. The remaining 22 studies were either of weak quality or included confounding promotions.

#### Retail promotions: consumption outcomes

Three studies^37,40,43^ of weak quality assessed the association between in-premise marketing and consumption-related outcomes in a retail environment (supporting information, S4). Two of these^37,40^ yielded evidence that in-premise marketing was associated with an increase in the consumption of targeted fruit and vegetables, with the third^43^ providing inconsistent evidence of effect direction. No evidence of statistical significance was observed in these studies, and all three included confounding promotions.

#### Retail promotions: attention outcomes

One weak quality study without confounding promotions^57^ found that colourful, branded shelf labelling promoting carbonated soft drinks was significantly associated with an increase in consumers’ visual attention, compared to standard unbranded shelf labelling.

#### Retail promotions: Body Mass Index

One moderate quality study without confounding promotions^59^ found that participants’ exposure to end-of-aisle displays, special floor displays and checkout displays was associated with an increase in body mass index. Inconsistent evidence of statistical significance was found, with this association significant for exposures to sugar sweetened beverage displays and displays of foods high in solid fats and added sugars when combined with sugar sweetened beverage displays. This association was non-significant for displays of foods high in solid fats and added sugars alone or fruit, vegetable and whole grain displays.

#### Retail promotions: dietary quality

Three studies were included which examine the association between in-premise marketing and indices of diet quality.^43,44,59^ One moderate quality study^59^ without confounding promotions did not report effect direction, but found no significant association between end-of-aisle display, special floor display or checkout display exposure and an index of healthy eating. The remaining two studies^43,44^ are both methodologically weak and included confounding promotions. One of these^44^ observed that exposure to entrance and checkout displays was associated with a significant improvement in diet quality. The other^43^ found that exposure to checkout placement, in-premise signage and leaflets was not consistently associated with dietary quality, with no evidence of statistical significance observed.

### Out-of-Home Promotions

Overall, evidence of in-premise marketing’s positive influence on consumer behaviour (e.g., increased sales, consumption or preferences of targeted items) was seen in 20 of 28 observed outcomes (across 24 studies, see supporting information, S5) in an out-of-home setting, with this association being significant in 9 studies. Two studies found evidence of promotions being associated with a significant negative influence on consumer behaviour (reduced sales and consumer preference), and six found inconsistent evidence with respect to effect direction.

### Out-of-Home Promotions: Sales

Eighteen studies examined the effect of in-premise marketing on sales in an out-of-home setting (supporting information, S5). Overall 16 of these (89%) found that positional promotions and/or in-premise advertising are associated with an increase in sales of promoted foods. This association was significant in eight studies, with inconsistent evidence of significance found in three studies. One methodologically weak study^69^ observed that checkout placement was associated with a significant decrease in sales of promoted foods.

The strongest evidence of the impact of in-premise marketing on sales in an out-of-home setting comes from one methodologically strong study^67^, which does not include confounding promotions. This study observed that verbal point-of-sale prompts were associated with significant increases in sales of pancakes, orange juice and fruit juice. Two additional studies of moderate methodological quality were conducted without confounding promotions.^42,53^ These found that verbal prompts^53^ and in-premise posters and table tents (self-standing folded flyers)^42^ were associated with increases in sales of promoted items. This association was significant in one study^53^ with no null hypothesis significance testing conducted in the other.^42^

#### Out-of-Home Promotions: Consumption

Three methodologically weak studies examined the effect of in-premise marketing on consumption outcomes in an out-of-home environment.^32,43,51^ Two of these were conducted in the absence of confounding promotions.^32,51^ One of these^51^ provided evidence that in-premise advertising was associated with increased consumption, with the remaining studies^32,43^ both providing inconsistent evidence of the direction of effect. No evidence of statistical significance was found across these three studies.

#### Out-of-Home Promotions: Consumer preference

Six studies assessed the association between in-premise advertising and consumer preferences in an out-of-home setting, all of which were conducted in the absence of confounding promotions. Overall, three of these^47,51,81^ found evidence in-premise advertising (verbal prompts, text and images on menus) are associated with increased consumption of promoted items, however this was only statistically significant in one study.^81^ One methodologically weak study^31^ found that menu imagery was associated with a significant reduction in consumer preferences of burgers and salad. The remaining two studies^41,56^ found inconsistent evidence of effect direction, with no indication of statistical significance.

The best available evidence of the impact of in-premise marketing on consumer preference in an out-of-home setting comes from one methodologically moderate study.^47^ This found evidence that verbal prompts are associated with a non-significant increase in consumer preferences for vegetarian meatballs.

#### Out-of-Home Promotions: Dietary quality

One methodologically weak study conducted in the presence of confounding promotions^43^ found that exposure to checkout placement, in-premise signage and leaflets was not consistently associated with dietary quality, with no evidence of statistical significance observed.

### Online Promotions

Overall, evidence of promotions’ positive influence on consumer behaviour (increased sales and increased consumer preference) was seen in two (both consumer preference) of six observed outcomes (across 6 studies^35,50,62,75,77,82^, see supporting information, S6) in an online setting, with this association being significant in one (supporting information, S6). Three studies found inconsistent evidence of effect direction,^35,50,77^ and one^82^ found that online promotion was associated with a negative influence on consumer behaviour (reduced sales).

#### Online Promotions: Sales

Two studies examined the impact of online promotions on sales.^35,82^ One methodologically moderate study^35^ found that positioning items on the first page of an online shopping environment (in the presence of other confounding promotions) was inconsistently associated with sales of targeted foods. One methodologically weak study^82^ without confounding promotions found that text prompts advising other consumers’ typical purchase quantity were associated with a reduction in sales of cookies, with inconsistent evidence of statistical significance.

#### Online Promotions: Consumer preference

Four studies examined the impact of online promotions on consumer preference, all in isolation from confounding promotions.^50,62,75,77^ Two methodologically moderate studies^62,75^ found that pop-up prompts offering healthier alternatives to chosen items were associated with an increase in the likelihood that promoted healthier items were chosen. This association was significant in one study.^62^ Two methodologically weak studies found inconsistent evidence of effect direction,^50,77^ with one finding inconsistent evidence of statistical significance.^50^

## Discussion

This systematic review investigated the impact of positional promotions and in-premise advertising implemented in retail, out-of-home and online environments upon consumer behaviour and diet-related outcomes. Three research questions were posed, relating to the quantity, quality and observations of the available evidence, in order to inform the development of public policy.

Overall, 62 research studies were identified, published over the period 1988 to 2020, with the majority of these being published in 2011 or later (84%). The included literature was not equally distributed across the settings considered in this review, with substantially fewer studies examining the impact of promotions in an online environment. Whilst a greater quantity of evidence focused on retail and out-of-home settings, this largely examined associations between promotions and sales outcomes, with a paucity of evidence on other outcome categories (particularly consumption, attention and health outcomes). As such, conclusions that can be made about the impact of promotions in an online environment, and on non-sales outcomes are limited. It was also not possible to identify any potential differential impact of in-premise marketing across demographic groups, due to sparse reporting of outcomes disaggregated by these characteristics.

The majority of the included literature was methodologically poor, with even the identified randomised controlled studies largely being at high risk of bias owing to potential selection biases, sparse information on randomisation processes, an inability to blind participants or researchers to exposure and high withdrawal rates. Whilst experimental studies typically offer better quality evidence, their utility in this context may be inappropriate due to inherent limitations. Three of the four studies identified as being methodologically strong utilised an observational design.^70,73,74^ These observational studies achieve this rating according to the EPHPP tool,^27^ in part, owing to their use of large datasets pertaining to real-world customers, which helps circumvent potential issues around blinding and selection biases. The quality of these studies was further enhanced by the use of statistical adjustment of potential confounders such as demographic characteristics, time/season, and confounding promotional strategies such as price.

Four methodologically strong studies^67,70,73,74^ represent the best available evidence of associations with consumer outcomes, across retail and out of home settings. These predominantly found evidence that in-premise marketing was associated with increased sales of targeted items (or reduced sales where such promotions are prohibited). These studies are strengthened by their examination of in-premise marketing in isolation from confounding promotions (e.g., price or availability) which could otherwise be responsible for driving observed associations.

Further to these methodologically strong studies, the ten identified as being of moderate quality and without confounding promotions yielded similar findings. For ten of the 11 outcomes presented in these studies (6 relating to sales outcomes,^42,46,53,68,79,90^ 3 consumer preference,^47,62,75^ 1 BMI,^59^ 1 diet quality^59^), evidence was found that is consistent with in-premise marketing being successful in impacting individuals’ responses to targeted foods. These findings span the three settings considered in this review, with six outcomes examined in retail settings, three out-of-home and two in online environments. Evidence of statistical significance was found for five of these 11 outcomes.

The lower quality studies that were included tended to be less consistent with respect to the observed direction of effect, which may reflect that the reported research tended to utilise smaller samples of individuals, transactions and/or premises. However across the entirety of the included studies, few found internally consistent (i.e., with 70% or more of relevant results in the same direction) evidence that in-premise marketing is associated with reduction in sales, attention, consumption or preferences, with only four methodologically weak studies observing this.^31,58,69,82^ One further weak study^86^ found that a displacement intervention actively promoting fruit and vegetables was associated with a non-significant increase in confectionary sales.

Reporting of quantitative metrics including effect size and confidence intervals was sparse across the included literature. This limits the possibility to make formal judgements of the consistency, precision and magnitude of observed effects. As such, it is impossible to formally apply GRADE^92^ criteria to assess the certainty of this body of evidence. The lack of sufficient information meant it was also not possible to formally assess publication bias. Very few of the included studies presented an assessment of any potential dose-response relationship between degrees of exposure to in-premise marketing and consumer outcomes, with the vast majority merely comparing the presence versus the absence of a promotional strategy. One exception in this regard is a study^59^ which estimated the effect of cumulative individual exposures to displays of sugar sweetened beverages, discretionary and healthy foods on BMI and dietary quality. Given the high risk of bias for the majority of the included literature, the certainty of this body of evidence as a whole is likely to be low or very low. A greater degree of certainty, however, may apply to a subset of the included literature, particularly the four studies identified as being methodologically strong.

A limitation of the included literature is the lack of evidence across all of the setting and outcome permutations considered here. The best available evidence pertains to sales data, which are only a proximal target of policy intervention in this area, with improvements in consumption and ultimately health status being preferential targets. Evidence comparing in-store purchases against 24 hour dietary recalls, however, suggests that sales outcomes do constitute a reasonable proxy of consumption behaviour.^93^ This may be particularly true for foods that are hedonistically pleasurable and/or convenient to consume on the go, like many discretionary products.

A-priori sample size calculations were conducted in less than a quarter of the included studies, with fewer still reporting that an adequate sample size had been achieved (10 studies). This may mean that many of the included studies are underpowered to detect statistically significant associations between in-premise marketing and the outcomes considered. Whilst statistical significance was not used to form conclusions based on the included evidence, the utilised data on effect direction may also be compromised in terms of its precision and representativeness with smaller samples. It was also rare for the included research to make appropriate adjustments when conducting multiple tests of statistical significance, which increases the likelihood of making conclusions on the basis of spurious associations. A further limitation of the included evidence is that prospective trial registration is particularly rare. This can allow researchers’ conscious or unconscious biases to manipulate the design, implementation and analyses of research studies, reducing the confidence that can be placed in the reported findings.^94^

Strengths of the methodology used to conduct this review include dual screening at all stages, and extraction of data and quality appraisal in duplicate, which increases accuracy and reduces bias in these stages. Prospective registration of the review protocol limits the extent to which the authors of this review could have introduced biases by making ad-hoc changes to the scope, methodology, synthesis or dissemination. Bibliographic databases were searched from their inception, minimising the potential for relevant literature to be omitted. Adherence to PRISMA,^24^ SWiM^25^ and Cochrane^29^ guidance optimises the transparency, repeatability and rigour of this review.

Limitations of this review include that the specific content of in-premise advertisements was not assessed (e.g., dish of the day promotion, price marking, social norms, health messages). It may be the case that specific messages, or particular features of in-premise advertisements such as colours/images/sounds are more effective in highlighting promoted products. It is also possible that there are differences between individuals in terms of perception and interpretation of these characteristics, as is the case for product packaging.^95,96^ The precise positioning of placement interventions was also not considered, although it may be the case that displays in different locations have more reliable or larger effects on consumer behaviour. The interactive nature of the marketing mix was also not a focus of the present review. This constitutes a limitation as the impacts of a product’s packaging, price, placement and promotion on consumer behaviour are potentially synergistic. For example a pricing intervention might be expected to have a greater effect if it is both in a prominent position and accompanied by in-store advertising to highlight its price. Including only English-language publications is a further limitation as any relevant studies published in other languages have not been incorporated into the synthesis presented here.

The vote counting methodology used was based on effect direction alone^28,29^ and provides no information on the magnitude or precision of observed effects. Therefore it may be the case that whilst the best available evidence points to a positive influence of in-premise marketing, that these may be small effects with limited impact on population health. Contrary to this speculation, however, one of the included methodologically strong studies explicitly addresses this issue,^73^ and presents effect size in terms of an equivalent price change. This study found that end-of-aisle displays of non-alcoholic beverages are associated with an increase in sales of these products equivalent to a reduction in price of between 22% and 62%.

It is key for further research in this area to address some of the chief methodological limitations of the extant evidence base, and to fill the knowledge gaps identified by the present review. Investigations are particularly needed to determine the role and impact of non-price promotions in online shopping environments due to the increasing importance of this purchasing setting. Research is also required which goes beyond sales data outcomes and examines associations between exposures to in-premise marketing and consumption of targeted items, and ultimately health endpoints. A salient absence in the included literature is investigation of potential demographic differences in susceptibility and exposure to in-premise marketing. Analysis of the impact of in-premise marketing by age, sex and socioeconomic status would inform efforts to narrow health inequalities.

Given the inherent difficulty with which randomised controlled trials can be conducted within commercial food environments, natural experiments of real-world policy changes likely constitute an optimal research design in these settings. With appropriate linkage of datasets, routinely collected information such as hospitalisation records may be used to determine the association between exposure to in-premise marketing and non-communicable disease morbidity and mortality. There would be value in supplementing such investigations with qualitative research and analysis of process variables such as consumers’ attitudes towards targeted products and the promotions themselves to understand the mechanisms behind any changes in behaviour. In order to address the specific methodological issues in the extant literature, future research in this context should also prioritise prospective registration, appropriate sample size calculations and transparency of reporting.

### Implications for policy

The included literature provides a basis for appropriate authorities to consider acting to restrict in-premise marketing of unhealthy foods, and encourage the marketing of healthier products. When considering the stronger available evidence, the observed promotional strategies appear to be successful in influencing consumer behaviour. Very little evidence was retrieved that in-premise marketing was associated with reductions in consumer response, with these findings restricted to methodologically weak studies. Where such promotions are targeting less healthy foods, this may lead to deterioration of population health, and vice-versa with foods that are more nutritionally beneficial.

One methodologically strong study^74^ found evidence which is of direct relevance to the development of public policy as it examines the impact of a restrictive intervention to remove discretionary foods from supermarket checkout displays, mirroring the potential form of national regulation in this area. Using a nationally representative sample of around 30,000 UK shoppers, this study found evidence that sales of discretionary foods were subject to an immediate (4 weeks after implementation) and sustained (12 months after implementation) reduction within supermarkets implementing policies to remove discretionary products from checkout displays, compared to supermarkets that did not change their checkout display policy. One further moderate quality study^46^ offers evidence that the prominent positioning of healthier foods is associated with a non-significant reduction in sales of sugar sweetened beverages. This represents a further direct test of a potential policy option to discourage population intake of less healthy products.

## Conclusion

Whilst the methodological quality of the included English-language evidence was predominantly poor, the best available evidence points to non-price in-premise marketing’s efficacy in eliciting the intended behavioural responses. The better-quality evidence predominantly pertains to sales outcomes in retail and out-of-home settings, with a relative paucity of evidence identified on the role of point-of-purchase promotions in online settings and for non-sales outcomes.

## Supporting information

Table 1. Inclusion and exclusion criteria

Figure 1. PRISMA flow diagram

## Data Availability

Data extracted from articles included in this systematic review are presented as supplementary information.

## Acknowledgements

The authors would like to thank the following members of the review advisory group for their role in helping establish the scope and rationale of the present review and for helping interpret its findings: Professor Linda Bauld, Col Baird, Dr Nathan Critchlow, Dr Kathryn Waite, Emma Riches, Rebecca Craig, Dr Margaret Callaghan, Lynda Brown.

## Abbreviations

HFSS: (foods) high in fat, sugar or salt
PRISMA: Preferred Reporting Items for Systematic Reviews and Meta-Analyses
SWiM: Synthesis Without Meta-analysis
BMI: body mass index (kg/m^2^)
EPHPP: Effective Public Health Practice Project
UK: United Kingdom
GRADE: Grading of Recommendations, Assessment, Development and Evaluation

## References

1. Global Strategy on Diet, Physical Activity and Health. World Health Organization; 2004. https://www.who.int/dietphysicalactivity/strategy/eb11344/strategy_english_web.pdf. Accessed March 31, 2021.

2. Set of recommendations on the marketing of’ ‘foods and non-alcoholic beverages to children.. World Health Organization; 2010. https://apps.who.int/iris/bitstream/handle/10665/44416/9789241500210_eng.pdf?sequence=1. Accessed March 31, 2021.

3. Blüher M. Obesity: global epidemiology and pathogenesis. Nat Rev Endocrinol. 2019;15(5):288–298. doi:10.1038/s41574-019-0176-8

4. Garrido-Miguel M, Cavero-Redondo I, Álvarez-Bueno C, et al. Prevalence and Trends of Overweight and Obesity in European Children From 1999 to 2016: A Systematic Review and Meta–analysis. JAMA Pediatr. August 2019:e192430. doi:10.1001/jamapediatrics.2019.2430

5. Cohen DA, Lesser LI. Obesity prevention at the point of purchase. Obes Rev. 2016;17(5):389–396. doi:10.1111/obr.12387

6. Cairns GA. Is the Emperor Naked? Rethinking approaches to responsible food marketing policy and research. March 2016. https://dspace.stir.ac.uk/handle/1893/23933#.YGSGga9KiMp. Accessed March 31, 2021.

7. Cairns G, Angus K, Hastings G, Caraher M. Systematic reviews of the evidence on the nature, extent and effects of food marketing to children. A retrospective summary. Appetite. 2013;62:209–215. doi:10.1016/j.appet.2012.04.017

8. Thornton LE, Cameron AJ, McNaughton SA, et al. Does the availability of snack foods in supermarkets vary internationally? Int J Behav Nutr Phys Act. 2013;10:56. doi:10.1186/1479-5868-10-56

9. Basch CH, Kernan WD, Menafro A. Presence of candy and snack food at checkout in chain stores: results of a pilot study. J Community Health. 2016;41(5):1090–1093. doi:10.1007/s10900-016-0193-7

10. Chandon P, Wansink B. Does food marketing need to make us fat? A review and solutions. Nutr Rev. 2012;70(10):571–593. doi:10.1111/j.1753-4887.2012.00518.x

11. Andreyeva T, Long MW, Brownell KD. The impact of food prices on consumption: a systematic review of research on the price elasticity of demand for food. Am J Public Health. 2010;100(2):216–222. doi:10.2105/AJPH.2008.151415

12. Cairns G. The’ ‘Impact of Food and Drink Marketing on Scotland’s Children and Young People: A report on the results of questions about exposure and purchase responses included in IPSOS - Mori’s 2014 Young People in Scotland Survey. University of Stirling, Institute of Social Marketing; 2015. https://www.stir.ac.uk/media/stirling/services/faculties/sport-and-health-sciences/documents/Impact-of-Food-and-Drink-on-Scotlands-Young---Sept-15.pdf. Accessed March 31, 2021.

13. Shaw SC, Ntani G, Baird J, Vogel CA. A systematic review of the influences of food store product placement on dietary-related outcomes. Nutr Rev. 2020;78(12):1030–1045. doi:10.1093/nutrit/nuaa024

14. Backholer K, Sacks G, Cameron AJ. Food and beverage price promotions: an untapped policy target for improving population diets and health. Curr Nutr Rep. 2019;8(3):250–255. doi:10.1007/s13668-019-00287-z

15. Kaur A, Lewis T, Lipkova V, et al. A systematic review, and meta-analysis, examining the prevalence of price promotions on foods and whether they are more likely to be found on less-healthy foods. Public Health Nutr. 2020;23(8):1281–1296. doi:10.1017/S1368980019004129

16. Sugar Reduction: Achieving the 20% A technical report outlining progress to date, guidelines for industry, 2015 baseline levels in key foods and next steps. Public Health England; 2017. https://assets.publishing.service.gov.uk/government/uploads/system/uploads/attachment_data/file/604336/Sugar_reduction_achieving_the_20_.pdf. Accessed March 31, 2021.

17. Setterfield L, Eunson J, Murray L. Marketing strategies used within premises by out of home businesses. Food Standards Scotland; 2017. https://www.foodstandards.gov.scot/downloads/Ipsos_Mori_-_marketing_strategies.pdf. Accessed March 31, 2021.

18. Fleischhacker SE, Evenson KR, Rodriguez DA, Ammerman AS. A systematic review of fast food access studies. Obes Rev. 2011;12(5):e460–71. doi:10.1111/j.1467-789X.2010.00715.x

19. Jilcott Pitts SB, Ng SW, Blitstein JL, Gustafson A, Niculescu M. Online grocery shopping: promise and pitfalls for healthier food and beverage purchases. Public Health Nutr. 2018;21(18):3360–3376. doi:10.1017/S1368980018002409

20. Obesity Action Scotland. Obesity Action Scotland | Healthy Weight For All - Survey of Food and Drink Promotions in an Online Retail Environment. https://obesityactionscotland.org/publications/reports/survey-of-food-and-drink-promotions-in-an-online-retail-environment/. Accessed March 31, 2021.

21. Gao X, Shi X, Guo H, Liu Y. To buy or not buy food online: The impact of the COVID-19 epidemic on the adoption of e-commerce in China. PLoS One. 2020;15(8):e0237900. doi:10.1371/journal.pone.0237900

22. Chang H, Meyerhoefer CD. COVID -19 and the Demand for Online Food Shopping Services: Empirical Evidence from Taiwan. Am J Agric Econ. 2021;103(2):448–465. doi:10.1111/ajae.12170

23. United Nations Conference on Trade and Development. COVID-19 has changed online shopping forever, survey shows | UNCTAD. https://unctad.org/news/covid-19-has-changed-online-shopping-forever-survey-shows. Accessed March 31, 2021.

24. Liberati A, Altman DG, Tetzlaff J, et al. The PRISMA statement for reporting systematic reviews and meta-analyses of studies that evaluate healthcare interventions: explanation and elaboration. BMJ. 2009;339:b2700. doi:10.1136/bmj.b2700

25. Campbell M, McKenzie JE, Sowden A, et al. Synthesis without meta-analysis (SWiM) in systematic reviews: reporting guideline. BMJ. 2020;368:n6890. doi:10.1136/bmj.l6890

26. Covidence - Better systematic review management. https://www.covidence.org/. Accessed March 31, 2021.

27. Effective Public Healthcare Panacea Project. Quality Assessment Tool for Quantitative Studies. https://www.ephpp.ca/quality-assessment-tool-for-quantitative-studies/. Accessed March 31, 2021.

28. Boon MH, Thomson H. The effect direction plot revisited: Application of the 2019 Cochrane Handbook guidance on alternative synthesis methods. Res Synth Methods. 2021;12(1):29–33. doi:10.1002/jrsm.1458

29. Cochrane Collaboration. Chapter 12: Synthesizing and presenting findings using other methods | Cochrane Training. https://training.cochrane.org/handbook/current/chapter-12. Accessed January 25, 2021.

30. Ayala GX, Castro IA, Pickrel JL, et al. A cluster randomized trial to promote healthy menu items for children: the kids’ choice restaurant program. Int J Environ Res Public Health. 2017;14(12). doi:10.3390/ijerph14121494

31. Gala P, Rippé CB, Dubinsky AJ, Favia MJ. Effects of menu calorie information and product image on millenials’ purchase intention. Marketing Management Journal. 2018;28(2):127–144.

32. Anzman-Frasca S, Braun AC, Ehrenberg S, et al. Effects of a randomized intervention promoting healthy children’s meals on children’s ordering and dietary intake in a quick-service restaurant. Physiol Behav. 2018;192:109–117. doi:10.1016/j.physbeh.2018.01.022

33. Dannefer R, Williams DA, Baronberg S, Silver L. Healthy bodegas: increasing and promoting healthy foods at corner stores in New York City. Am J Public Health. 2012;102(10):e27–31. doi:10.2105/AJPH.2011.300615

34. French SA, Jeffery RW, Story M, et al. Pricing and promotion effects on low-fat vending snack purchases: the CHIPS Study. Am J Public Health. 2001;91(1):112–117. doi:10.2105/ajph.91.1.112

35. Delaney T, Wyse R, Yoong SL, et al. Cluster randomized controlled trial of a consumer behavior intervention to improve healthy food purchases from online canteens. Am J Clin Nutr. 2017;106(5):1311–1320. doi:10.3945/ajcn.117.158329

36. Foster GD, Karpyn A, Wojtanowski AC, et al. Placement and promotion strategies to increase sales of healthier products in supermarkets in low-income, ethnically diverse neighborhoods: a randomized controlled trial. Am J Clin Nutr. 2014;99(6):1359–1368. doi:10.3945/ajcn.113.075572

37. Kristal AR, Goldenhar L, Muldoon J, Morton RF. Evaluation of a supermarket intervention to increase consumption of fruits and vegetables. Am J Health Promot. 1997;11(6):422–425. doi:10.4278/0890-1171-11.6.422

38. Fiske A, Cullen KW. Effects of promotional materials on vending sales of low-fat items in teachers’ lounges. J Am Diet Assoc. 2004;104(1):90–93. doi:10.1016/j.jada.2003.10.011

39. Hua SV, Kimmel L, Van Emmenes M, et al. Health promotion and healthier products increase vending purchases: A randomized factorial trial. J Acad Nutr Diet. 2017;117(7):1057–1065. doi:10.1016/j.jand.2016.12.006

40. Ayala GX, Baquero B, Laraia BA, Ji M, Linnan L. Efficacy of a store-based environmental change intervention compared with a delayed treatment control condition on store customers’ intake of fruits and vegetables. Public Health Nutr. 2013;16(11):1953–1960. doi:10.1017/S1368980013000955

41. Bacon L, Krpan D. (Not) Eating for the environment: The impact of restaurant menu design on vegetarian food choice. Appetite. 2018;125:190–200. doi:10.1016/j.appet.2018.02.006

42. Wagner JL, Winett RA. Prompting one low-fat, high-fiber selection in a fast-food restaurant. J Appl Behav Anal. 1988;21(2):179–185. doi:10.1901/jaba.1988.21-179

43. Trude ACB, Surkan PJ, Cheskin LJ, Gittelsohn J. A multilevel, multicomponent childhood obesity prevention group-randomized controlled trial improves healthier food purchasing and reduces sweet-snack consumption among low-income African-American youth. Nutr J. 2018;17(1):96. doi:10.1186/s12937-018-0406-2

44. Baird J, Crozier SR, Penn-Newman D, Cooper C, Vogel C. RF27 How does changing the placement of food products in supermarkets influence customers’ diets? In: Oral presentations. BMJ Publishing Group Ltd; 2018:A55.2-A56. doi:10.1136/jech-2018-SSMabstracts.115

45. Sanchez-Flack J, Pickrel JL, Belch G, et al. Examination of the Relationship between In-Store Environmental Factors and Fruit and Vegetable Purchasing among Hispanics. Int J Environ Res Public Health. 2017;14(11). doi:10.3390/ijerph14111305

46. Huitink M, Poelman MP, Seidell JC, et al. Can unhealthy food purchases at checkout counters be discouraged by introducing healthier snacks? A real-life experiment in supermarkets in deprived urban areas in the Netherlands. BMC Public Health. 2020;20(1):542. doi:10.1186/s12889-020-08608-6

47. Dos Santos Q, Perez-Cueto FJA, Rodrigues VM, et al. Impact of a nudging intervention and factors associated with vegetable dish choice among European adolescents. Eur J Nutr. 2020;59(1):231–247. doi:10.1007/s00394-019-01903-y

48. Pharis ML, Colby L, Wagner A, Mallya G. Sales of healthy snacks and beverages following the implementation of healthy vending standards in City of Philadelphia vending machines. Public Health Nutr. 2018;21(2):339–345. doi:10.1017/S1368980017001914

49. Collins EIM, Thomas JM, Robinson E, et al. Two observational studies examining the effect of a social norm and a health message on the purchase of vegetables in student canteen settings. Appetite. 2019;132:122–130. doi:10.1016/j.appet.2018.09.024

50. Breugelmans E, Campo K, Gijsbrechts E. Shelf sequence and proximity effects on online grocery choices. Mark Lett. 2007;18(1-2):117–133. doi:10.1007/s11002-006-9002-x

51. Schwartz MB. The influence of a verbal prompt on school lunch fruit consumption: a pilot study. Int J Behav Nutr Phys Act. 2007;4:6. doi:10.1186/1479-5868-4-6

52. Lee-Kwan SH, Bleich SN, Kim H, Colantuoni E, Gittelsohn J. Environmental Intervention in Carryout Restaurants Increases Sales of Healthy Menu Items in a Low-Income Urban Setting. Am J Health Promot. 2015;29(6):357–364. doi:10.4278/ajhp.130805-QUAN-408

53. Saulais L, Massey C, Perez-Cueto FJA, et al. When are “Dish of the Day” nudges most effective to increase vegetable selection? Food Policy. 2019;85:15–27. doi:10.1016/j.foodpol.2019.04.003

54. Sigurdsson V, Larsen NM, Gunnarsson D. Healthy food products at the point of purchase: An in-store experimental analysis. J Appl Behav Anal. 2014;47(1):151–154. doi:10.1002/jaba.91

55. Salmon SJ, De Vet E, Adriaanse MA, Fennis BM, Veltkamp M, De Ridder DTD. Social proof in the supermarket: Promoting healthy choices under low self-control conditions. Food Qual Prefer. 2015;45:113–120. doi:10.1016/j.foodqual.2015.06.004

56. Nazlan NH, Tanford S, Raab C, Choi C (CB). The influence of scarcity cues and price bundling on menu item selection. Journal of Foodservice Business Research. 2018;21(4):420–439. doi:10.1080/15378020.2018.1440129

57. Hurley RA, Rice JC, Koefelda J, Congdon R, Ouzts A. The role of secondary packaging on brand awareness: analysis of 2 L carbonated soft drinks in reusable shells using eye tracking technology. Packag Technol Sci. 2017;30(11):711–722. doi:10.1002/pts.2316

58. Budd N, Jeffries JK, Jones-Smith J, Kharmats A, McDermott AY, Gittelsohn J. Store-directed price promotions and communications strategies improve healthier food supply and demand: Impact results from a randomized controlled, Baltimore City store-intervention trial. Public Health Nutr. 2017;20(18):3349–3359. doi:10.1017/S1368980017000064

59. Cohen DA, Collins R, Hunter G, Ghosh-Dastidar B, Dubowitz T. Store impulse marketing strategies and body mass index. Am J Public Health. 2015;105(7):1446–1452. doi:10.2105/AJPH.2014.302220

60. Gustafson A, Ng SW, Jilcott Pitts S. The association between the “Plate it Up Kentucky” supermarket intervention and changes in grocery shopping practices among rural residents. Transl Behav Med. 2019;9(5):865–874. doi:10.1093/tbm/ibz064

61. Broers VJV, Van den Broucke S, Taverne C, Luminet O. Default-name and tasting nudges increase salsify soup choice without increasing overall soup choice. Appetite. 2019;138:204–214. doi:10.1016/j.appet.2019.03.027

62. Koutoukidis DA, Jebb SA, Ordóñez-Mena JM, et al. Prominent positioning and food swaps are effective interventions to reduce the saturated fat content of the shopping basket in an experimental online supermarket: a randomized controlled trial. Int J Behav Nutr Phys Act. 2019;16(1):50. doi:10.1186/s12966-019-0810-9

63. Mistura M, Fetterly N, Rhodes RE, Tomlin D, Naylor P-J. Examining the Efficacy of a “Feasible” Nudge Intervention to Increase the Purchase of Vegetables by First Year University Students (17-19 Years of Age) in British Columbia: A Pilot Study. Nutrients. 2019;11(8). doi:10.3390/nu11081786

64. Chapman LE, Sadeghzadeh C, Koutlas M, Zimmer C, De Marco M. Evaluation of three behavioural economics “nudges” on grocery and convenience store sales of promoted nutritious foods. Public Health Nutr. 2019;22(17):3250–3260. doi:10.1017/S1368980019001794

65. Lopez NV, Folta SC, Glenn ME, Lynskey VM, Patel AA, Anzman-Frasca S. Promoting healthier children’s meals at quick-service and full-service restaurants: Results from a pilot and feasibility study. Appetite. 2017;117:91–97. doi:10.1016/j.appet.2017.06.015

66. Lawman HG, Vander Veur S, Mallya G, et al. Changes in quantity, spending, and nutritional characteristics of adult, adolescent and child urban corner store purchases after an environmental intervention. Prev Med. 2015;74:81–85. doi:10.1016/j.ypmed.2014.12.003

67. van Kleef E, van den Broek O, van Trijp HCM. Exploiting the Spur of the Moment to Enhance Healthy Consumption: Verbal Prompting to Increase Fruit Choices in a Self-Service Restaurant. Appl Psychol Health Well Being. 2015;7(2):149–166. doi:10.1111/aphw.12042

68. Payne C, Niculescu M. Can healthy checkout end-caps improve targeted fruit and vegetable purchases? Evidence from grocery and SNAP participant purchases. Food Policy. 2018;79:318–323. doi:10.1016/j.foodpol.2018.03.002

69. Chapman K, Ogden J. Nudging Customers towards Healthier Choices: An Intervention in the University Canteen. J Food Res. 2012;1(2). doi:10.5539/jfr.v1n2p13

70. Daunfeldt S-O, Rudholm N. Does shelf-labeling of organic foods increase sales? Results from a natural experiment. Journal of Retailing and Consumer Services. 2014;21(5):804–811. doi:10.1016/j.jretconser.2014.06.009

71. Winkler LL, Christensen U, Glümer C, et al. Substituting sugar confectionery with fruit and healthy snacks at checkout - a win-win strategy for consumers and food stores? a study on consumer attitudes and sales effects of a healthy supermarket intervention. BMC Public Health. 2016;16(1):1184. doi:10.1186/s12889-016-3849-4

72. Wolfenden L, Kingsland M, Rowland BC, et al. Improving availability, promotion and purchase of fruit and vegetable and non sugar-sweetened drink products at community sporting clubs: a randomised trial. Int J Behav Nutr Phys Act. 2015;12(1):193. doi:10.1186/s12966-015-0193-5

73. Nakamura R, Pechey R, Suhrcke M, Jebb SA, Marteau TM. Sales impact of displaying alcoholic and non-alcoholic beverages in end-of-aisle locations: an observational study. Soc Sci Med. 2014;108:68–73. doi:10.1016/j.socscimed.2014.02.032

74. Ejlerskov KT, Sharp SJ, Stead M, Adamson AJ, White M, Adams J. Supermarket policies on less-healthy food at checkouts: Natural experimental evaluation using interrupted time series analyses of purchases. PLoS Med. 2018;15(12):e1002712. doi:10.1371/journal.pmed.1002712

75. Forwood SE, Ahern AL, Marteau TM, Jebb SA. Offering within-category food swaps to reduce energy density of food purchases: a study using an experimental online supermarket. Int J Behav Nutr Phys Act. 2015;12:85. doi:10.1186/s12966-015-0241-1

76. Caspi CE, Lenk K, Pelletier JE, et al. Association between store food environment and customer purchases in small grocery stores, gas-marts, pharmacies and dollar stores. Int J Behav Nutr Phys Act. 2017;14(1):76. doi:10.1186/s12966-017-0531-x

77. Nederkoorn C. Effects of sales promotions, weight status, and impulsivity on purchases in a supermarket. Obesity (Silver Spring). 2014;22(5):E2–5. doi:10.1002/oby.20621

78. Gamburzew A, Darcel N, Gazan R, et al. In-store marketing of inexpensive foods with good nutritional quality in disadvantaged neighborhoods: increased awareness, understanding, and purchasing. Int J Behav Nutr Phys Act. 2016;13(1):104. doi:10.1186/s12966-016-0427-1

79. Van Gestel LC, Kroese FM, De Ridder DTD. Nudging at the checkout counter - A longitudinal study of the effect of a food repositioning nudge on healthy food choice. Psychol Health. 2018;33(6):800–809. doi:10.1080/08870446.2017.1416116

80. Ebster C, Wagner U, Valis S. The effectiveness of verbal prompts on sales. Journal of Retailing and Consumer Services. 2006;13(3):169–176. doi:10.1016/j.jretconser.2005.08.003

81. Hou Y, Yang W, Sun Y. Do pictures help? The effects of pictures and food names on menu evaluations. Int J Hosp Manag. 2017;60:94–103. doi:10.1016/j.ijhm.2016.10.008

82. Jove” Hou J. Can Interface Cues Nudge Modeling of Food Consumption? Experiments on a Food-Ordering Website. J Comput-Mediat Comm. 2017;22(4):196–214. doi:10.1111/jcc4.12190

83. Fitzgerald CM, Kannan S, Sheldon S, Eagle KA. Effect of a promotional campaign on heart-healthy menu choices in community restaurants. J Am Diet Assoc. 2004;104(3):429–432. doi:10.1016/j.jada.2003.12.019

84. Garrido-Morgado Á, González-Benito Ó. Merchandising at the point of sale: differential effect of end of aisle and islands. BRQ Business Research Quarterly. 2015;18(1):57–67. doi:10.1016/j.brq.2013.11.004

85. Sigurdsson V, Larsen NM, Gunnarsson D. An in-store experimental analysis of consumers’ selection of fruits and vegetables. The Service Industries Journal. 2011;31(15):2587–2602. doi:10.1080/02642069.2011.531126

86. Buscher LA, Martin KA, Crocker S. Point-of-purchase messages framed in terms of cost, convenience, taste, and energy improve healthful snack selection in a college foodservice setting. J Am Diet Assoc. 2001;101(8):909–913. doi:10.1016/S0002-8223(01)00223-1

87. Ensaff H, Homer M, Sahota P, Braybrook D, Coan S, McLeod H. Food Choice Architecture: An Intervention in a Secondary School and its Impact on Students’ Plant-based Food Choices. Nutrients. 2015;7(6):4426–4437. doi:10.3390/nu7064426

88. Toft U, Winkler LL, Mikkelsen BE, Bloch P, Glümer C. Discounts on fruit and vegetables combined with a space management intervention increased sales in supermarkets. Eur J Clin Nutr. 2017;71(4):476–480. doi:10.1038/ejcn.2016.272

89. Paine-Andrews A, Francisco VT. Health marketing in the supermarket: Using prompting, product sampling, and price reduction to… Health Marketing Quarterly. 1996;14(2):85.

90. L Harris J, Webb V, J Sacco S, L Pomeranz J. Marketing to children in supermarkets: an opportunity for public policy to improve children’s diets. Int J Environ Res Public Health. 2020;17(4). doi:10.3390/ijerph17041284

91. Cheung TTL, Gillebaart M, Kroese FM, Marchiori D, Fennis BM, De Ridder DTD. Cueing healthier alternatives for take-away: a field experiment on the effects of (disclosing) three nudges on food choices. BMC Public Health. 2019;19(1):974. doi:10.1186/s12889-019-7323-y

92. Guyatt GH, Oxman AD, Vist GE, et al. GRADE: an emerging consensus on rating quality of evidence and strength of recommendations. BMJ. 2008;336(7650):924–926. doi:10.1136/bmj.39489.470347.AD

93. Appelhans BM, French SA, Tangney CC, Powell LM, Wang Y. To what extent do food purchases reflect shoppers’ diet quality and nutrient intake? Int J Behav Nutr Phys Act. 2017;14(1):46. doi:10.1186/s12966-017-0502-2

94. Wicherts JM, Veldkamp CLS, Augusteijn HEM, Bakker M, van Aert RCM, van Assen MALM. Degrees of Freedom in Planning, Running, Analyzing, and Reporting Psychological Studies: A Checklist to Avoid p-Hacking. Front Psychol. 2016;7:1832. doi:10.3389/fpsyg.2016.01832

95. Mai R, Symmank C, Seeberg-Elverfeldt B. Light and pale colors in food packaging: when does this package cue signal superior healthiness or inferior tastiness? Journal of Retailing. 2016;92(4):426–444. doi:10.1016/j.jretai.2016.08.002

96. Bialkova S, Sasse L, Fenko A. The role of nutrition labels and advertising claims in altering consumers’ evaluation and choice. Appetite. 2016;96:38–46. doi:10.1016/j.appet.2015.08.030

