## Supporting information (S1-S6) for "The Impact of Non-Price In-premise Marketing on Food and Beverage Purchasing and Consumer Behaviour: A Systematic Review"

^1^ Place and Wellbeing Directorate, Public Health Scotland.

^2^ MRC/CSO Social and Public Health Sciences Unit, University of Glasgow

^3^ Institute for Social Marketing and Health, University of Stirling 
 

**Supporting information S1.** Search strategy

**Database: Business Source Complete**

**Interface/Platform:** EBSCOhost Research Databases

**Limiters:** Scholarly (Peer Reviewed) Journals; Publication Type: Academic Journal; Language: English

| **Search ID no.** | **Search terms** |
| --- | --- |
|  | ***Context/Setting terms*** |
| S1 | TI bodega# OR AB bodega# |
| S2 | TI c-store# OR AB c-store# |
| S3 | TI ( café# OR cafe# ) OR AB ( café# OR cafe# ) |
| S4 | TI ( caféteria# OR cafeteria# ) OR AB ( caféteria# OR cafeteria# ) |
| S5 | TI canteen# OR AB canteen# |
| S6 | TI cinema# OR AB cinema# |
| S7 | TI click-and-collect OR AB click-and-collect |
| S8 | TI ( consum* W1 (food* OR snack* OR meal* OR drink* OR beverage# OR junk* OR "fast-food#" OR "calorie-dense" OR supper* OR dinner* OR breakfast* OR lunch*) ) OR AB ( consum* W1 (food* OR snack* OR meal* OR drink* OR beverage# OR junk* OR "fast-food#" OR "calorie-dense" OR supper* OR dinner* OR breakfast* OR lunch*) ) |
| S9 | TI ( convenience W2 (retail* OR shop# OR store#) ) OR AB ( convenience W2 (retail* OR shop# OR store#) ) |
| S10 | TI ( corner W2 (retail* OR shop# OR store#) ) OR AB ( corner W2 (retail* OR shop# OR store#) ) |
| S11 | TI ( coffee W2 (retail* OR shop# OR store#) ) OR AB ( coffee W2 (retail* OR shop# OR store#) ) |
| S12 | TI ( grocery W2 (retail* OR shop# OR store#) ) OR AB ( grocery W2 (retail* OR shop# OR store#) ) |
| S13 | TI ( sandwich W2 (retail* OR shop# OR store#) ) OR AB ( sandwich W2 (retail* OR shop# OR store#) ) |
| S14 | TI diner# OR AB diner# |
| S15 | TI drive-in# OR AB drive-in# |
| S16 | TI "eating place#" OR AB "eating place#" |
| S17 | TI "fast food" OR AB "fast food" |
| S18 | TI "fast service" OR AB "fast service" |
| S19 | TI ( food-court# OR foodcourt# ) OR AB ( food-court# OR foodcourt# ) |
| S20 | TI "food environment" OR AB "food environment" |
| S21 | TI "food on-the-go" OR AB "food on-the-go" |
| S22 | TI ( food N2 (van# OR truck#) ) OR AB ( food N2 (van# OR truck#) ) |
| S23 | TI food N2 stall# OR AB food N2 stall# |
| S24 | TI food N2 concession# OR AB food N2 concession# |
| S25 | TI food N2 outlet# OR AB food N2 outlet# |
| S26 | TI ( food N2 (delivery OR deliveries) ) OR AB ( food N2 (delivery OR deliveries) ) |
| S27 | TI food N2 store# OR AB food N2 store# |
| S28 | TI "food market#" OR AB "food market#" |
| S29 | TI hypermarket# OR AB hypermarket# |
| S30 | TI instore# OR AB instore# |
| S31 | TI "internal environment#" OR AB "internal environment#" |
| S32 | TI kiosk* OR AB kiosk* |
| S33 | TI mall# OR AB mall# |
| S34 | TI movie W1 theat* OR AB movie W1 theat* |
| S35 | TI newsagent# OR AB newsagent# |
| S36 | TI "out-of-home" OR AB "out-of-home" |
| S37 | TI "out-of-house" OR AB "out-of-house" |
| S38 | TI pop-up# OR AB pop-up# |
| S39 | TI precinct# OR AB precinct# |
| S40 | TI "quick service" OR AB "quick service" |
| S41 | TI restaurant# OR AB restaurant# |
| S42 | TI ( (retail* OR shop# OR store#) W2 environment ) OR AB ( (retail* OR shop# OR store#) W2 environment ) |
| S43 | TI "self-service*" OR AB "self-service*" |
| S44 | TI supermarket# OR AB supermarket# |
| S45 | TI ( take-away# OR takeaway# ) OR AB ( take-away# OR takeaway# ) |
| S46 | TI ( take-out# OR takeout# ) OR AB ( take-out# OR takeout# ) |
| S47 | TI vending OR AB vending |
| S48 | TI vendor* OR AB vendor* |
| S49 | TI ( customer# W2 (food* OR snack* OR meal* OR drink* OR beverage# OR junk* OR "fast-food#" OR "calorie-dense" OR supper* OR dinner* OR breakfast* OR lunch*) ) OR AB ( customer# W2 (food* OR snack* OR meal* OR drink* OR beverage# OR junk* OR "fast-food#" OR "calorie-dense" OR supper* OR dinner* OR breakfast* OR lunch*) ) |
| S50 | TI ( commerc* N2 (food* OR snack* OR meal* OR drink* OR beverage# OR junk* OR "fast-food#" OR "calorie-dense" OR supper* OR dinner* OR breakfast* OR lunch*) ) OR AB ( commerc* N2 (food* OR snack* OR meal* OR drink* OR beverage# OR junk* OR "fast-food#" OR "calorie-dense" OR supper* OR dinner* OR breakfast* OR lunch*) ) |
| S51 | TI ( premise# W2 (food* OR snack* OR meal* OR drink* OR beverage# OR junk* OR "fast-food#" OR "calorie-dense" OR supper* OR dinner* OR breakfast* OR lunch*) ) OR AB ( premise# W2 (food* OR snack* OR meal* OR drink* OR beverage# OR junk* OR "fast-food#" OR "calorie-dense" OR supper* OR dinner* OR breakfast* OR lunch*) ) |
| S52 | TI ( retail* W2 (food* OR snack* OR meal* OR drink* OR beverage# OR junk* OR "fast-food#" OR "calorie-dense" OR supper* OR dinner* OR breakfast* OR lunch*) ) OR AB ( retail* W2 (food* OR snack* OR meal* OR drink* OR beverage# OR junk* OR "fast-food#" OR "calorie-dense" OR supper* OR dinner* OR breakfast* OR lunch*) ) |
| S53 | TI ( sale# N2 (food* OR snack* OR meal* OR drink* OR beverage# OR junk* OR "fast-food#" OR "calorie-dense" OR supper* OR dinner* OR breakfast* OR lunch*) ) OR AB ( sale# N2 (food* OR snack* OR meal* OR drink* OR beverage# OR junk* OR "fast-food#" OR "calorie-dense" OR supper* OR dinner* OR breakfast* OR lunch*) ) |
| S54 | TI ( sell# N2 (food* OR snack* OR meal* OR drink* OR beverage# OR junk* OR "fast-food#" OR "calorie-dense" OR supper* OR dinner* OR breakfast* OR lunch*) ) OR AB ( sell# N2 (food* OR snack* OR meal* OR drink* OR beverage# OR junk* OR "fast-food#" OR "calorie-dense" OR supper* OR dinner* OR breakfast* OR lunch*) ) |
| S55 | TI ( app# N2 (delivery OR deliveries OR grocer* OR retailer# OR shop* OR store# OR supermarket*) ) OR AB ( app# N2 (delivery OR deliveries OR grocer* OR retailer# OR shop* OR store# OR supermarket*) ) |
| S56 | TI ( internet N2 (delivery OR deliveries OR grocer* OR retailer# OR shop* OR store# OR supermarket*) ) OR AB ( internet N2 (delivery OR deliveries OR grocer* OR retailer# OR shop* OR store# OR supermarket*) ) |
| S57 | TI ( mobile N2 (delivery OR deliveries OR grocer* OR retailer# OR shop* OR store# OR supermarket*) ) OR AB ( mobile N2 (delivery OR deliveries OR grocer* OR retailer# OR shop* OR store# OR supermarket*) ) |
| S58 | TI ( online N2 (delivery OR deliveries OR grocer* OR retailer# OR shop* OR store# OR supermarket*) ) OR AB ( online N2 (delivery OR deliveries OR grocer* OR retailer# OR shop* OR store# OR supermarket*) ) |
| S59 | TI ( smartphone# N2 (delivery OR deliveries OR grocer* OR retailer# OR shop* OR store# OR supermarket*) ) OR AB ( smartphone# N2 (delivery OR deliveries OR grocer* OR retailer# OR shop* OR store# OR supermarket*) ) |
| S60 | TI ( ("cell phone#" OR cellphone#) N2 (delivery OR deliveries OR grocer* OR retailer# OR shop* OR store# OR supermarket*) ) OR AB ( ("cell phone#" OR cellphone#) N2 (delivery OR deliveries OR grocer* OR retailer# OR shop* OR store# OR supermarket*) ) |
| S61 | TI ( web N2 (delivery OR deliveries OR grocer* OR retailer# OR shop* OR store# OR supermarket*) ) OR AB ( web N2 (delivery OR deliveries OR grocer* OR retailer# OR shop* OR store# OR supermarket*) ) |
| S62 | TI ( website# N2 (delivery OR deliveries OR grocer* OR retailer# OR shop* OR store# OR supermarket*) ) OR AB ( website# N2 (delivery OR deliveries OR grocer* OR retailer# OR shop* OR store# OR supermarket*) ) |
| S63 | S1 OR S2 OR S3 OR S4 OR S5 OR S6 OR S7 OR S8 OR S9 OR S10 OR S11 OR S12 OR S13 OR S14 OR S15 OR S16 OR S17 OR S18 OR S19 OR S20 OR S21 OR S22 OR S23 OR S24 OR S25 OR S26 OR S27 OR S28 OR S29 OR S30 OR S31 OR S32 OR S33 OR S34 OR S35 OR S36 OR S37 OR S38 OR S39 OR S40 OR S41 OR S42 OR S43 OR S44 OR S45 OR S46 OR S47 OR S48 OR S49 OR S50 OR S51 OR S52 OR S53 OR S54 OR S55 OR S56 OR S57 OR S58 OR S59 OR S60 OR S61 OR S62 |
|  | ***Intervention/Exposure (food) terms*** |
| S64 | TI ( food* OR snack* OR meal* OR drink* OR beverage# OR junk* OR "fast-food#" OR "calorie-dense" OR supper* OR dinner* OR breakfast* OR lunch* ) OR AB ( food* OR snack* OR meal* OR drink* OR beverage# OR junk* OR "fast-food#" OR "calorie-dense" OR supper* OR dinner* OR breakfast* OR lunch* ) |
|  | ***Intervention/Exposure (promotion) terms*** |
| S65 | TI advert* OR AB advert* |
| S66 | TI marketing OR AB marketing |
| S67 | TI promotion* OR AB promotion* |
| S68 | TI offer# OR AB offer# |
| S69 | TI sponsor* OR AB sponsor* |
| S70 | TI aisle* OR AB aisle* |
| S71 | TI ambient OR AB ambient |
| S72 | TI atmospheric# OR AB atmospheric# |
| S73 | TI banner# OR AB banner# |
| S74 | TI bin OR AB bin |
| S75 | TI cabinet* OR AB cabinet* |
| S76 | TI chiller* OR AB chiller* |
| S77 | TI display* OR AB display* |
| S78 | TI (endcap# OR end-cap#) OR AB (endcap# OR end-cap#) |
| S79 | TI gondola# OR AB gondola# |
| S80 | TI impuls* OR AB impuls* |
| S81 | TI instore# OR AB instore# |
| S82 | TI island OR AB island |
| S83 | TI merchandis* OR AB merchandis* |
| S84 | TI prompt* OR AB prompt* |
| S85 | TI screen# OR AB screen# |
| S86 | TI "shelf-edg*" OR AB "shelf-edg*" |
| S87 | TI ( "shelf-talker#" OR "shelftalker#" ) OR AB ( "shelf-talker#" OR "shelftalker#" ) |
| S88 | TI ( shelfspac* OR shelf-spac* ) OR AB ( shelfspac* OR shelf-spac* ) |
| S89 | TI sign# OR AB sign# |
| S90 | TI signage OR AB signage |
| S91 | TI ( activation# N2 (brand* OR store* OR in-store OR retail OR on-premise OR in-premise) ) OR AB ( activation# N2 (brand* OR store* OR in-store OR retail OR on-premise OR in-premise) ) |
| S92 | TI ( pop-up# OR popup# ) OR AB ( pop-up# OR popup# ) |
| S93 | TI freebie# OR AB freebie# |
| S94 | TI flyer# OR AB flyer# |
| S95 | TI leaflet# OR AB leaflet# |
| S96 | TI magazine# OR AB magazine# |
| S97 | TI menu# OR AB menu# |
| S98 | TI "recipe card#" OR AB "recipe card#" |
| S99 | TI "consumer-oriented" OR AB "consumer-oriented" |
| S100 | TI "customer-oriented" OR AB "customer-oriented" |
| S101 | TI "choice architect*" OR AB "choice architect*" |
| S102 | TI nudg* OR AB nudg* |
| S103 | TI layout# OR AB layout# |
| S104 | TI placement# OR AB placement# |
| S105 | TI "cash desk*" OR AB "cash desk*" |
| S106 | TI cashier* OR AB cashier* |
| S107 | TI "cash register*" OR AB "cash register*" |
| S108 | TI ("check-out#" OR checkout#) OR AB ("check-out#" OR checkout#) |
| S109 | TI point# N3 purchas* OR AB point# N3 purchas* |
| S110 | TI point# N3 sale* OR AB point# N3 sale* |
| S111 | TI point# N3 till* OR AB point# N3 till* |
| S112 | TI tillpoint# OR AB tillpoint# |
| S113 | TI celebrit* OR AB celebrit* |
| S114 | TI influencer# OR AB influencer# |
| S115 | TI selling OR AB selling |
| S116 | TI staff* N2 bonus* OR AB staff* N2 bonus* |
| S117 | TI staff* N2 incentiv* OR AB staff* N2 incentiv* |
| S118 | TI ( supersiz* OR "super-siz*" ) OR AB ( supersiz* OR "super-siz*" ) |
| S119 | TI ( upsell* OR "up-sell*" ) OR AB ( upsell* OR "up-sell*" ) |
| S120 | TI announc* OR AB announc* |
| S121 | TI ( loudspeaker* OR "loud-speaker*" ) OR AB ( loudspeaker* OR "loud-speaker*" ) |
| S122 | TI "public-address*" OR AB "public-address*" |
| S123 | TI radio OR AB radio |
| S124 | TI beacon# OR AB beacon# |
| S125 | TI "customer service#" OR AB "customer service#" |
| S126 | TI ( attention W5 (retail* OR shop# OR store# OR supermarket#) ) OR AB ( attention W5 (retail* OR shop# OR store# OR supermarket#) ) |
| S127 | TI ( design* W5 (retail* OR shop# OR store# OR supermarket#) ) OR AB ( design* W5 (retail* OR shop# OR store# OR supermarket#) ) |
| S128 | TI ( feature* W5 (retail* OR shop# OR store# OR supermarket#) ) OR AB ( feature* W5 (retail* OR shop# OR store# OR supermarket#) ) |
| S129 | TI ( highlight* W5 (retail* OR shop# OR store# OR supermarket#) ) OR AB ( highlight* W5 (retail* OR shop# OR store# OR supermarket#) ) |
| S130 | TI ( recommend* W2 (consumer* OR customer* OR staff* OR retail* OR shop# OR store# OR supermarket# ) OR AB ( recommend* W2 (consumer* OR customer* OR staff* OR retail* OR shop# OR store# OR supermarket# ) |
| S131 | S65 OR S66 OR S67 OR S68 OR S69 OR S70 OR S71 OR S72 OR S73 OR S74 OR S75 OR S76 OR S77 OR S78 OR S79 OR S80 OR S81 OR S82 OR S83 OR S84 OR S85 OR S86 OR S87 OR S88 OR S89 OR S90 OR S91 OR S92 OR S93 OR S94 OR S95 OR S96 OR S97 OR S98 OR S99 OR S100 OR S101 OR S102 OR S103 OR S104 OR S105 OR S106 OR S107 OR S108 OR S109 OR S110 OR S111 OR S112 OR S113 OR S114 OR S115 OR S116 OR S117 OR S118 OR S119 OR S120 OR S121 OR S122 OR S123 OR S124 OR S125 OR S126 OR S127 OR S128 OR S129 OR S130 |
| S132 | S63 AND S64 AND S131 |
|  | ***Intervention/Exposure (promotion and food) terms – using database index and no setting terms*** |
| S133 | DE "DISPLAY advertising" OR DE "DISPLAY of merchandise" OR DE "POINT-of-sale advertising" OR DE "RETAIL space allocation" |
| S134 | food* OR snack* OR meal* OR drink* OR beverageS OR junk* OR "fast-foodS" OR "calorie-dense" OR supper* OR dinner* OR breakfast* OR lunch* |
| S135 | S133 AND S134 |
|  | ***Combined results*** |
| S136 | S132 OR S135 |

**Databases: The Cochrane Database of Systematic Reviews; The Cochrane Central Register of Controlled Trials (CENTRAL)**

**Interface/Platform:** The Cochrane Library

**Limiters:** Title Abstract Keyword search

| **Search ID no.** | **Search terms** |
| --- | --- |
| #1 | (marketing OR advertising OR promote OR promotion) AND (retail* OR wholesaler OR (cash NEAR carry) OR shop* OR store* OR mall* OR "shopping NEAR centre" OR supermarket* OR hypermarket* OR grocer*) AND (food* OR drink* OR snack* OR beverage*) |

**Database: EconLit**

**Interface/Platform:** EBSCOhost Research Databases

**Limiters:** Language: English

| **Search ID no.** | **Search terms** |
| --- | --- |
| S1 | TI ( bodega# OR c-store# OR café# OR cafe# OR caféteria# OR cafeteria# OR canteen# OR cinema# OR click-and-collect OR diner# OR drive-in# OR "eating place#" OR "fast food" OR "fast service" OR food-court# OR foodcourt# OR "food environment" OR "food on-the-go" OR "food market#" OR hypermarket# OR instore# OR "internal environment#" OR kiosk* OR mall# OR (movie W1 theat*) OR newsagent# OR "out-of-home" OR "out-of-house" OR pop-up# OR precinct# OR "quick service" OR restaurant# OR "self-service*" OR supermarket# OR take-away# OR takeaway# OR take-out# OR takeout# OR vending OR vendor* ) OR AB ( bodega# OR c-store# OR café# OR cafe# OR caféteria# OR cafeteria# OR canteen# OR cinema# OR click-and-collect OR diner# OR drive-in# OR "eating place#" OR "fast food" OR "fast service" OR food-court# OR foodcourt# OR "food environment" OR "food on-the-go" OR "food market#" OR hypermarket# OR instore# OR "internal environment#" OR kiosk* OR mall# OR (movie W1 theat*) OR newsagent# OR "out-of-home" OR "out-of-house" OR pop-up# OR precinct# OR "quick service" OR restaurant# OR "self-service*" OR supermarket# OR take-away# OR takeaway# OR take-out# OR takeout# OR vending OR vendor* ) |
| S2 | TI ( consum* W1 (food* OR snack* OR meal* OR drink* OR beverage# OR junk* OR "fast-food#" OR "calorie-dense" OR supper* OR dinner* OR breakfast* OR lunch*) ) OR AB ( consum* W1 (food* OR snack* OR meal* OR drink* OR beverage# OR junk* OR "fast-food#" OR "calorie-dense" OR supper* OR dinner* OR breakfast* OR lunch*) ) |
| S3 | TI ( (convenience OR corner OR coffee OR grocery OR sandwich) W2 (retail* OR shop# OR store#) ) OR AB ( (convenience OR corner OR coffee OR grocery OR sandwich) W2 (retail* OR shop# OR store#) ) |
| S4 | TI ( food N2 (concession# OR delivery OR deliveries OR outlet# OR stall# OR store# OR van# OR truck#) ) OR AB ( food N2 (concession# OR delivery OR deliveries OR outlet# OR stall# OR store# OR van# OR truck#) ) |
| S5 | TI ( (retail* OR shop# OR store#) W2 environment ) OR AB ( (retail* OR shop# OR store#) W2 environment ) |
| S6 | TI ( (customer# OR premise# OR retail*) W2 (food* OR snack* OR meal* OR drink* OR beverage# OR junk* OR "fast-food#" OR "calorie-dense" OR supper* OR dinner* OR breakfast* OR lunch*) ) OR AB ( (customer# OR premise# OR retail*) W2 (food* OR snack* OR meal* OR drink* OR beverage# OR junk* OR "fast-food#" OR "calorie-dense" OR supper* OR dinner* OR breakfast* OR lunch*) ) |
| S7 | TI ( (commerc* OR sale# OR sell#) N2 (food* OR snack* OR meal* OR drink* OR beverage# OR junk* OR "fast-food#" OR "calorie-dense" OR supper* OR dinner* OR breakfast* OR lunch*) ) OR AB ( (commerc* OR sale# OR sell#) N2 (food* OR snack* OR meal* OR drink* OR beverage# OR junk* OR "fast-food#" OR "calorie-dense" OR supper* OR dinner* OR breakfast* OR lunch*) ) |
| S8 | TI ( (app# OR "cell phone#" OR cellphone# OR internet OR mobile OR online OR smartphone# OR web OR website#) N2 (delivery OR deliveries OR grocer* OR retailer# OR shop* OR store# OR supermarket*) ) OR AB ( (app# OR "cell phone#" OR cellphone# OR internet OR mobile OR online OR smartphone# OR web OR website#) N2 (delivery OR deliveries OR grocer* OR retailer# OR shop* OR store# OR supermarket*) ) |
| S9 | S1 OR S2 OR S3 OR S4 OR S5 OR S6 OR S7 OR S8 |
| S10 | TI ( food* OR snack* OR meal* OR drink* OR beverage# OR junk* OR "fast-food#" OR "calorie-dense" OR supper* OR dinner* OR breakfast* OR lunch* ) OR AB ( food* OR snack* OR meal* OR drink* OR beverage# OR junk* OR "fast-food#" OR "calorie-dense" OR supper* OR dinner* OR breakfast* OR lunch* ) |
| S11 | TI ( advert* OR marketing OR promotion* OR offer# OR sponsor* OR aisle* OR ambient OR atmospheric# OR banner# OR bin OR cabinet* OR chiller* OR display* OR endcap# OR end-cap# OR gondola# OR impuls* OR instore# OR island OR merchandis* OR prompt* OR screen# OR "shelf-edg*" OR "shelf-talker#" OR "shelftalker#" OR shelfspac* OR shelf-spac* OR sign# OR signage OR pop-up# OR popup# OR freebie# OR flyer# OR leaflet# OR magazine# OR menu# OR "recipe card#" OR "consumer-oriented" OR "customer-oriented" OR "choice architect*" OR nudg* OR layout# OR placement# OR "cash desk*" OR cashier* OR "cash register*" OR "check-out#" OR checkout# OR tillpoint# OR celebrit* OR influencer# OR selling OR supersiz* OR "super-siz*" OR upsell* OR "up-sell*" OR announc* OR loudspeaker* OR "loud-speaker*" OR "public-address*" OR radio OR beacon# OR "customer service#" ) OR AB ( advert* OR marketing OR promotion* OR offer# OR sponsor* OR aisle* OR ambient OR atmospheric# OR banner# OR bin OR cabinet* OR chiller* OR display* OR endcap# OR end-cap# OR gondola# OR impuls* OR instore# OR island OR merchandis* OR prompt* OR screen# OR "shelf-edg*" OR "shelf-talker#" OR "shelftalker#" OR shelfspac* OR shelf-spac* OR sign# OR signage OR pop-up# OR popup# OR freebie# OR flyer# OR leaflet# OR magazine# OR menu# OR "recipe card#" OR "consumer-oriented" OR "customer-oriented" OR "choice architect*" OR nudg* OR layout# OR placement# OR "cash desk*" OR cashier* OR "cash register*" OR "check-out#" OR checkout# OR tillpoint# OR celebrit* OR influencer# OR selling OR supersiz* OR "super-siz*" OR upsell* OR "up-sell*" OR announc* OR loudspeaker* OR "loud-speaker*" OR "public-address*" OR radio OR beacon# OR "customer service#" ) |
| S12 | TI ( activation# N2 (brand* OR store* OR in-store OR retail OR on-premise OR in-premise) ) OR AB ( activation# N2 (brand* OR store* OR in-store OR retail OR on-premise OR in-premise) ) |
| S13 | TI ( point# N3 (purchas* OR sale* OR till*) ) OR AB ( point# N3 (purchas* OR sale* OR till*) ) |
| S14 | TI ( staff* N2 (bonus* OR incentiv*) ) OR AB ( staff* N2 (bonus* OR incentiv*) ) |
| S15 | TI ( (attention OR design* OR feature* OR highlight*) W5 (retail* OR shop# OR store# OR supermarket#) ) OR AB ( (attention OR design* OR feature* OR highlight*) W5 (retail* OR shop# OR store# OR supermarket#) ) |
| S16 | TI ( recommend* W2 (consumer* OR customer* OR staff* OR retail* OR shop# OR store# OR supermarket#) ) OR AB ( recommend* W2 (consumer* OR customer* OR staff* OR retail* OR shop# OR store# OR supermarket#) ) |
| S17 | S11 OR S12 OR S13 OR S14 OR S15 OR S16 |
| S18 | S9 AND S10 AND S17 |
| S19 | food* OR snack* OR meal* OR drink* OR beverage# OR junk* OR "fast-food#" OR "calorie-dense" OR supper* OR dinner* OR breakfast* OR lunch* |
| S20 | ZU "consumer economics expenditure patterns and consumption of specific items" OR ZU "consumer motivation. brand preference" OR ZU "consumption of other individual goods OR services" OR ((ZU "retail food" or ZU "retail and wholesale trade; e-commerce" or ZU "retailing") AND (ZU "advertising" OR ZU "marketing" OR ZU "marketing and advertising" OR ZU "marketing and advertising: general")) |
| S21 | S19 AND S20 |
| S22 | S18 OR S21 |
| S23 | ( S18 OR S21 ) AND ( ZL "english" OR ZL "englishenglish" ) |

**Database: Emerald Insight**

**Interface/Platform:** EBSCO Discovery Service

**Limiters:** None

| **Search ID no.** | **Search terms** |
| --- | --- |
| S1 | AB ( (food* OR snack* OR meal* OR drink* OR beverage* OR junk* OR "fast-food" OR "calorie-dense" OR supper* OR dinner* OR breakfast* OR lunch*) ) OR TI ( (food* OR snack* OR meal* OR drink* OR beverage* OR junk* OR "fast-food" OR "calorie-dense" OR supper* OR dinner* OR breakfast* OR lunch*) ) |
| S2 | AB ( (recommend* N1 (consumer* OR customer* OR staff* OR retail* OR shop* OR store* OR supermarket*) ) OR AB ( (attention OR design* OR feature* OR highlight*) N2 (retail* OR shop* OR store* OR supermarket*) ) OR AB ( (staff*) N1 (bonus* OR incentiv*) ) OR AB ( (point* N1 (purchas* OR sale* OR till*)) ) OR AB ( (activation* N1 (brand* OR store* OR in-store* OR retail OR on-premise* OR in-premise*)) ) OR AB ( (aisle* OR ambient OR atmospheric* OR banner* OR bin OR cabinet* OR chiller* OR endcap$ OR end-cap* OR gondola* OR instore* OR island OR merchandis* OR screen* OR "shelf-edg*" OR "shelf-talker*" OR "shelftalker*" OR shelfspac* OR "shelf-spac*" OR sign* OR signage OR pop-up* OR popup* OR freebie* OR flyer* OR leaflet* OR magazine* OR "recipe card*" OR "consumer-oriented" OR "customer-oriented" OR "choice architect*" OR nudg* OR layout* OR placement* OR "cash desk*" OR cashier* OR "cash register*" OR "check-out*" OR checkout* OR tillpoint* OR celebrit* OR influencer* OR selling OR supersiz* OR "super-siz*" OR upsell* OR "up-sell*" OR announc* OR loudspeaker* OR "loud-speaker*" OR "public-address*" OR "radio" OR beacon*) ) OR AB ( (advert* OR marketing OR promotion* OR offer* OR sponsor* OR display* OR menu* OR impuls* OR prompt*) N2 (food* OR snack* OR meal* OR drink* OR beverage$ OR junk* OR "fast-food*" OR "calorie-dense" OR supper* OR dinner* OR breakfast* OR lunch*) ) |
| S3 | AB ( (commerc* OR customer* OR premise* OR retail* OR sale* OR sell*) N2 (food* OR snack* OR meal* OR drink* OR beverage* OR junk* OR "fast-food*" OR "calorie-dense" OR supper* OR dinner* OR breakfast* OR lunch*)) ) OR AB ( (food N1 (concession* OR deliveries OR delivery OR outlet* OR stall* OR store* OR truck* OR van*)) ) OR AB ( (convenience OR corner OR coffee OR environment OR grocery OR sandwich) N1 (retail* OR shop* OR store*)) ) OR AB ( (consum* N1 (food* OR snack* OR meal* OR drink* OR beverage* OR junk OR "fast-food*" OR "calorie-dense" OR supper* OR dinner* OR breakfast* OR lunch*)) ) OR AB ( (bodega* OR c-store* OR café* OR café* OR cafeteria* OR cafeteria* OR canteen* OR cinema* OR click-and-collect OR diner* OR drive-in* OR "eating place*" OR "fast food" OR "fast service" OR food-court* OR foodcourt* OR "food environment" OR "food on-the-go" OR "food market*" OR hypermarket* OR "in-store*" OR instore* OR "in-premise*" OR "internal environment*" OR kiosk* OR mall* OR movie) N1 (theat* OR newsagent$ OR "on-premise*" OR "out-of-home" OR "out-of-house" OR pop-up* OR precinct$ OR "quick service" OR restaurant* OR "self-service*" OR supermarket* OR take-away* OR takeaway* OR take-out* OR takeout* OR vending OR vendor*)) ) |
| S4 | S1 AND S2 AND S3 |

**Database: APA PsycINFO**

**Interface/Platform:** EBSCOhost Research Databases

**Limiters:** Language: English

| **Search ID no.** | **Search terms** |
| --- | --- |
| S1 | TI ( bodega# OR c-store# OR café# OR cafe# OR caféteria# OR cafeteria# OR canteen# OR cinema# OR click-and-collect OR diner# OR drive-in# OR "eating place#" OR "fast food" OR "fast service" OR food-court# OR foodcourt# OR "food environment" OR "food on-the-go" OR "food market#" OR hypermarket# OR instore# OR "internal environment#" OR kiosk* OR mall# OR (movie W1 theat*) OR newsagent# OR "out-of-home" OR "out-of-house" OR pop-up# OR precinct# OR "quick service" OR restaurant# OR "self-service*" OR supermarket# OR take-away# OR takeaway# OR take-out# OR takeout# OR vending OR vendor* ) OR AB ( bodega# OR c-store# OR café# OR cafe# OR caféteria# OR cafeteria# OR canteen# OR cinema# OR click-and-collect OR diner# OR drive-in# OR "eating place#" OR "fast food" OR "fast service" OR food-court# OR foodcourt# OR "food environment" OR "food on-the-go" OR "food market#" OR hypermarket# OR instore# OR "internal environment#" OR kiosk* OR mall# OR (movie W1 theat*) OR newsagent# OR "out-of-home" OR "out-of-house" OR pop-up# OR precinct# OR "quick service" OR restaurant# OR "self-service*" OR supermarket# OR take-away# OR takeaway# OR take-out# OR takeout# OR vending OR vendor* ) |
| S2 | TI ( consum* W1 (food* OR snack* OR meal* OR drink* OR beverage# OR junk* OR "fast-food#" OR "calorie-dense" OR supper* OR dinner* OR breakfast* OR lunch*) ) OR AB ( consum* W1 (food* OR snack* OR meal* OR drink* OR beverage# OR junk* OR "fast-food#" OR "calorie-dense" OR supper* OR dinner* OR breakfast* OR lunch*) ) |
| S3 | TI ( (convenience OR corner OR coffee OR grocery OR sandwich) W2 (retail* OR shop# OR store#) ) OR AB ( (convenience OR corner OR coffee OR grocery OR sandwich) W2 (retail* OR shop# OR store#) ) |
| S4 | TI ( food N2 (concession# OR delivery OR deliveries OR outlet# OR stall# OR store# OR van# OR truck#) ) OR AB ( food N2 (concession# OR delivery OR deliveries OR outlet# OR stall# OR store# OR van# OR truck#) ) |
| S5 | TI ( (retail* OR shop# OR store#) W2 environment ) OR AB ( (retail* OR shop# OR store#) W2 environment ) |
| S6 | TI ( (customer# OR premise# OR retail*) W2 (food* OR snack* OR meal* OR drink* OR beverage# OR junk* OR "fast-food#" OR "calorie-dense" OR supper* OR dinner* OR breakfast* OR lunch*) ) OR AB ( (customer# OR premise# OR retail*) W2 (food* OR snack* OR meal* OR drink* OR beverage# OR junk* OR "fast-food#" OR "calorie-dense" OR supper* OR dinner* OR breakfast* OR lunch*) ) |
| S7 | TI ( (commerc* OR sale# OR sell#) N2 (food* OR snack* OR meal* OR drink* OR beverage# OR junk* OR "fast-food#" OR "calorie-dense" OR supper* OR dinner* OR breakfast* OR lunch*) ) OR AB ( (commerc* OR sale# OR sell#) N2 (food* OR snack* OR meal* OR drink* OR beverage# OR junk* OR "fast-food#" OR "calorie-dense" OR supper* OR dinner* OR breakfast* OR lunch*) ) |
| S8 | TI ( (app# OR "cell phone#" OR cellphone# OR internet OR mobile OR online OR smartphone# OR web OR website#) N2 (delivery OR deliveries OR grocer* OR retailer# OR shop* OR store# OR supermarket*) ) OR AB ( (app# OR "cell phone#" OR cellphone# OR internet OR mobile OR online OR smartphone# OR web OR website#) N2 (delivery OR deliveries OR grocer* OR retailer# OR shop* OR store# OR supermarket*) ) |
| S9 | S1 OR S2 OR S3 OR S4 OR S5 OR S6 OR S7 OR S8 |
| S10 | TI ( food* OR snack* OR meal* OR drink* OR beverage# OR junk* OR "fast-food#" OR "calorie-dense" OR supper* OR dinner* OR breakfast* OR lunch* ) OR AB ( food* OR snack* OR meal* OR drink* OR beverage# OR junk* OR "fast-food#" OR "calorie-dense" OR supper* OR dinner* OR breakfast* OR lunch* ) |
| S11 | TI ( advert* OR marketing OR promotion* OR offer# OR sponsor* OR aisle* OR ambient OR atmospheric# OR banner# OR bin OR cabinet* OR chiller* OR display* OR endcap# OR end-cap# OR gondola# OR impuls* OR instore# OR island OR merchandis* OR prompt* OR screen# OR "shelf-edg*" OR "shelf-talker#" OR "shelftalker#" OR shelfspac* OR shelf-spac* OR sign# OR signage OR pop-up# OR popup# OR freebie# OR flyer# OR leaflet# OR magazine# OR menu# OR "recipe card#" OR "consumer-oriented" OR "customer-oriented" OR "choice architect*" OR nudg* OR layout# OR placement# OR "cash desk*" OR cashier* OR "cash register*" OR "check-out#" OR checkout# OR tillpoint# OR celebrit* OR influencer# OR selling OR supersiz* OR "super-siz*" OR upsell* OR "up-sell*" OR announc* OR loudspeaker* OR "loud-speaker*" OR "public-address*" OR radio OR beacon# OR "customer service#" ) OR AB ( advert* OR marketing OR promotion* OR offer# OR sponsor* OR aisle* OR ambient OR atmospheric# OR banner# OR bin OR cabinet* OR chiller* OR display* OR endcap# OR end-cap# OR gondola# OR impuls* OR instore# OR island OR merchandis* OR prompt* OR screen# OR "shelf-edg*" OR "shelf-talker#" OR "shelftalker#" OR shelfspac* OR shelf-spac* OR sign# OR signage OR pop-up# OR popup# OR freebie# OR flyer# OR leaflet# OR magazine# OR menu# OR "recipe card#" OR "consumer-oriented" OR "customer-oriented" OR "choice architect*" OR nudg* OR layout# OR placement# OR "cash desk*" OR cashier* OR "cash register*" OR "check-out#" OR checkout# OR tillpoint# OR celebrit* OR influencer# OR selling OR supersiz* OR "super-siz*" OR upsell* OR "up-sell*" OR announc* OR loudspeaker* OR "loud-speaker*" OR "public-address*" OR radio OR beacon# OR "customer service#" ) |
| S12 | TI ( activation# N2 (brand* OR store* OR in-store OR retail OR on-premise OR in-premise) ) OR AB ( activation# N2 (brand* OR store* OR in-store OR retail OR on-premise OR in-premise) ) |
| S13 | TI ( point# N3 (purchas* OR sale* OR till*) ) OR AB ( point# N3 (purchas* OR sale* OR till*) ) |
| S14 | TI ( staff* N2 (bonus* OR incentiv*) ) OR AB ( staff* N2 (bonus* OR incentiv*) ) |
| S15 | TI ( (attention OR design* OR feature* OR highlight*) W5 (retail* OR shop# OR store# OR supermarket#) ) OR AB ( (attention OR design* OR feature* OR highlight*) W5 (retail* OR shop# OR store# OR supermarket#) ) |
| S16 | TI ( recommend* W2 (consumer* OR customer* OR staff* OR retail* OR shop# OR store# OR supermarket#) ) OR AB ( recommend* W2 (consumer* OR customer* OR staff* OR retail* OR shop# OR store# OR supermarket#) ) |
| S17 | S11 OR S12 OR S13 OR S14 OR S15 OR S16 |
| S18 | S9 AND S10 AND S17 |
| S19 | (DE "Shopping" OR DE "Retailing" OR DE "Consumer Behavior" OR DE "Consumer Research") AND (DE "Advertising" OR DE "Marketing" OR DE "Digital Marketing" OR DE "Visual Tracking" OR DE "Contextual Cues" OR DE "Visual Attention") |
| S20 | food* OR snack* OR meal* OR drink* OR beverage# OR junk* OR "fast-food#" OR "calorie-dense" OR supper* OR dinner* OR breakfast* OR lunch* |
| S21 | S19 AND S20 |
| S22 | S18 OR S21 |

**Database: Web of Science Core Collection**

**Indices:** Science Citation Index Expanded (SCI-EXPANDED) – 1900-present; Social Sciences Citation Index (SSCI) – 1900-present; Emerging Sources Citation Index (ESCI) – 2005-present

**Interface/Platform:** Web of Science, Clarivate Analytics

**Limiters:** Language: English

| **Search ID no.** | **Search terms** |
| --- | --- |
| #1 | (TS=(bodega$  OR c-store$  OR café$  OR cafe$  OR caféteria$  OR cafeteria$  OR canteen$  OR cinema$  OR click-and-collect  OR diner$  OR drive-in$  OR "eating place$"  OR "fast food"  OR "fast service"  OR food-court$  OR foodcourt$  OR "food environment"  OR "food on-the-go"  OR "food market$"  OR hypermarket$  OR "in-store$"  OR instore$  OR "in-premise$"  OR "internal environment$"  OR kiosk*  OR mall$  OR movie NEAR/0 theat*  OR newsagent$  OR "on-premise$"  OR "out-of-home"  OR "out-of-house"  OR pop-up$  OR precinct$  OR "quick service"  OR restaurant$  OR "self-service*"  OR supermarket$  OR take-away$  OR takeaway$  OR take-out$  OR takeout$  OR vending  OR vendor*))  AND LANGUAGE: (English) |
| #2 | (TS=(consum* NEAR/1 (food*  OR snack*  OR meal*  OR drink*  OR beverage$  OR junk*  OR "fast-food$"  OR "calorie-dense"  OR supper*  OR dinner*  OR breakfast*  OR lunch*)))  AND LANGUAGE: (English) |
| #3 | (TS=((convenience  OR corner  OR coffee  OR environment  OR grocery  OR sandwich) NEAR/1 (retail*  OR shop$  OR store$)))  AND LANGUAGE: (English) |
| #4 | (TS=(food NEAR/1 (concession$  OR deliveries  OR delivery  OR outlet$  OR stall$  OR store$  OR truck$  OR van$)))  AND LANGUAGE: (English) |
| #5 | (TS=((commerc*  OR customer$  OR premise$  OR retail*  OR sale$  OR sell$) NEAR/2 (food*  OR snack*  OR meal*  OR drink*  OR beverage$  OR junk*  OR "fast-food$"  OR "calorie-dense"  OR supper*  OR dinner*  OR breakfast*  OR lunch*)))  AND LANGUAGE: (English) |
| #6 | (#5  OR #4  OR #3  OR #2  OR #1) |
| #7 | (TS=(food*  OR snack*  OR meal*  OR drink*  OR beverage$  OR junk*  OR "fast-food$"  OR "calorie-dense"  OR supper*  OR dinner*  OR breakfast*  OR lunch*))  AND LANGUAGE: (English) |
| #8 | (TS=((advert*  OR marketing  OR promotion*  OR offer$  OR sponsor*  OR display*  OR menu$  OR impuls*  OR prompt*)  NEAR (food*  OR snack*  OR meal*  OR drink*  OR beverage$  OR junk*  OR "fast-food$"  OR "calorie-dense"  OR supper*  OR dinner*  OR breakfast*  OR lunch*)))  AND LANGUAGE: (English) |
| #9 | (TS=(aisle*  OR ambient  OR atmospheric$  OR banner$  OR bin  OR cabinet*  OR chiller*  OR endcap$  OR end-cap$  OR gondola$  OR instore$  OR island  OR merchandis*  OR screen$  OR "shelf-edg*"  OR "shelf-talker$"  OR "shelftalker$"  OR shelfspac*  OR "shelf-spac*"  OR sign$  OR signage  OR pop-up$  OR popup$  OR freebie$  OR flyer$  OR leaflet$  OR magazine$  OR "recipe card$"  OR "consumer-oriented"  OR "customer-oriented"  OR "choice architect*"  OR nudg*  OR layout$  OR placement$  OR "cash desk*"  OR cashier*  OR "cash register*"  OR "check-out$"  OR checkout$  OR tillpoint$  OR celebrit*  OR influencer$  OR selling  OR supersiz*  OR "super-siz*"  OR upsell*  OR "up-sell*"  OR announc*  OR loudspeaker*  OR "loud-speaker*"  OR "public-address*"  OR "radio"  OR beacon$))  AND LANGUAGE: (English) |
| #10 | (TS=(activation$ NEAR/1 (brand*  OR store$  OR in-store$  OR retail  OR on-premise$  OR in-premise$)))  AND LANGUAGE: (English) |
| #11 | (TS=(point$ NEAR/1 (purchas*  OR sale*  OR till*)))  AND LANGUAGE: (English) |
| #12 | (TS=(staff* NEAR/1 (bonus*  OR incentiv*)))  AND LANGUAGE: (English) |
| #13 | (TS=((attention  OR design*  OR feature*  OR highlight*) NEAR/4 (retail*  OR shop$  OR store$  OR supermarket$)))  AND LANGUAGE: (English) |
| #14 | (TS=(recommend* NEAR/1 (consumer*  OR customer*  OR staff*  OR retail*  OR shop$  OR store$  OR supermarket$)))  AND LANGUAGE: (English) |
| #15 | (#14  OR #13  OR #12  OR #11  OR #10  OR #9  OR #8) |
| #16 | (#15  AND #7  AND #6) |

**Database: SAGE Business Cases**

**Interface/Platform:** SAGE Knowledge, <https://uk.sagepub.com/en-gb/eur/sage-business-cases>

**Search mode:** Advanced search

**Limiters:** by selected indexed subjects

| **Search for (full text):** | **Document Type:** | **Topic: Subject:** | **Refine Subjects:** |
| --- | --- | --- | --- |
| food | Cases | Business & Management | Consumer behavior |
| food | Cases | Business & Management | Consumer marketing |
| food | Cases | Business & Management | Retailing |
| food | Cases | Business & Management | Marketing Channels |
| food | Cases | Business & Management | Advertising & Promotion |
| food | Cases | Business & Management | Experience Marketing |
| food | Cases | Business & Management | Marketing Communications |
| food | Cases | Business & Management | New Product Design & Marketing |
| food | Cases | Business & Management | Product Management |
| food | Cases | Business & Management | Consumer Culture |

**Database: WARC (World Advertising Research Center)**

**Interface/Platform:** Ascential plc, <https://www.warc.com/aboutus.info>

**Limiters:** Articles and Research Papers

| **Browsed by** | **Limited by** | |
| --- | --- | --- |
| Topics > Media channels & touchpoints > **POP & in-store** | **Search in**  Articles  Research Papers | **Categories**  Food  Soft drinks |
| Topics > Media planning > **Context & position of advertising** | **Search in**  Articles  Research Papers | **Categories**  Food  Soft drinks |
| Topics > Media channels & touchpoints > **Events & experiential** | **Search in**  Articles  Research Papers | **Categories**  Food  Soft drinks |
| Topics > Media channels & touchpoints > **Mobile** | **Search in**  Articles  Research Papers | **Categories**  Food  Soft drinks |
| Topics > Media channels & touchpoints > **Online display** | **Search in**  Articles  Research Papers | **Categories**  Food  Soft drinks |
| Topics > Media channels & touchpoints > **Promotions** | **Search in**  Articles  Research Papers | **Categories**  Food  Soft drinks |
| Topics > Data, tech & innovation > **Retail & e-commerce** | **Search in**  Articles  Research Papers | **Categories**  Food  Soft drinks |
| Topics > Consumers & audiences > **Shopper insight** | **Search in**  Articles  Research Papers | **Categories**  Food  Soft drinks |
| Topics > Categories > **Food** | **Search in**  Research Papers | **Source**  *Int. Journal of Advertising*  *Int. Journal of Market Research*  *Int. Journal of Mobile Marketing*  *Journal of Advertising History*  *Journal of Advertising Research* |
| Topics > Categories > **Soft drinks** | **Search in**  Research Papers | **Source**  *Int. Journal of Advertising*  *Int. Journal of Market Research*  *Journal of Advertising Research* |

**Supporting information S2.** Descriptive characteristics of included studies of restrictive interventions, by quality and presence of confounding promotions

| **Outcome category** | **Author, publication year, country** | **Study design** | **Sample size** | **Interventions used (targeted products)** | **Confounding promotions** | **Direction of effect ^a^** | **Study quality** |
| --- | --- | --- | --- | --- | --- | --- | --- |
| Sales (‘take-home’ panel) | Ejlerskov, 2018, UK^74^ | Observational (12 months pre-, 12 months post-implementation) | 9 national supermarket chains, extrapolated panel of ~30,000 UK households) | Removal from checkout display (discretionary foods) | None | ▼ | Strong |
| Sales (electronically recorded) | Sigurdsson, 2014, Norway^54^ | Uncontrolled pre-post (3-4 weeks pre-, 9-10 weeks post implementation) | 1 convenience store, 1 supermarket (102,368 transactions) | Removal from checkout displays (confectionary, chewing gum) | None | ▽ ^b^ | Weak |
| Sales (electronically recorded) | Winkler, 2016, Denmark^71^ | Non-randomised, controlled study (4 weeks pre, 4 weeks post implementation) | 28 supermarkets, 24 control, 4 intervention) | Removal from checkout displays (confectionary) | None | ▽ | Weak |
| Diet quality (participant report) | Baird, 2018, UK^44^ | Non-randomised, controlled study (between-subjects pre-post implementation) | 138 individuals | Removal from checkout displays (confectionary) | Placement within supermarket | ▼^c^ | Weak |

^a^ Orientation of arrow denotes the direction of association between the promotion and the outcome (up: increase in 70% or more of relevant results; down: decrease in 70% or more of relevant results; horizontal: study provides inconsistent evidence of effect direction). The shading of the arrow denotes whether the association was statistically significant (filled: p≤.05 in 70% or more of relevant results; unfilled: p≥.05 in 70% or more of relevant results; grey: study provides inconsistent evidence of statistical significance).

^b^ No null-hypothesis significance testing was conducted for the comparison of interest.

^c^ An improvement in dietary quality is assumed to reflect a reduction in the consumption of the discretionary food items subject to a restrictive promotion.

**Supporting information S3**. Descriptive characteristics of included studies of displacement interventions, by quality and presence of confounding promotions

| **Outcome category** | **Author, publication year, country** | **Study design** | **Sample size** | **Promotions used ^a^** | **Confounding promotions** | **Direction of effect ^b^** | **Study quality** |
| --- | --- | --- | --- | --- | --- | --- | --- |
| Sales (electronically recorded) | Huitink, 2020, Netherlands^46^ | Non-randomised, controlled study (2 months pre-, 2 months post implementation) | 24 supermarkets (9 control, 15 intervention) | Checkout placement (promoting fruit, vegetables, water, healthier nut and cereal bars, effect upon confectionary measured) | None | ▽ | Moderate |
| Sales (sales receipts) | Gustafson, 2019, USA^60^ | Non-randomised, controlled study (repeated cross-section pre-, post-implementation) | 497 individuals, 16 supermarkets (6 control, 10 intervention) | End-of-aisle placement, recipe cards, grocery cart placards, branded banners (promoting healthy recipes including fruit and vegetables, effect upon sugar sweetened beverages measured) | Taste testing, price reductions | ▽ | Moderate |
| Sales (electronically and manually recorded) | Buscher, 2001, Canada^86^ | Uncontrolled pre-post (multiple baseline and intervention periods) | 1 cafeteria (~277,728 transactions) | In-premise posters (promoting fruit, vegetables, pretzels, yoghurt, effect upon confectionary measured) | None | △ | Weak |
| Sales (electronically recorded) | Cheung, 2019, Netherlands^91^ | Uncontrolled pre-post (1 week baseline, 2 weeks intervention) | 1 canteen (51,198 transactions) | Checkout placement, in-premise posters, presentation bin (promoting fruit, bread, effect upon confectionary and croissants measured) | None | ▼ | Weak |

^a^ Studies in this table examined the impact of promotions on non-promoted products (i.e., a displacement effect). Further details are given in the “Promotions used” column.

^b^ Orientation of arrow denotes the direction of association between the promotion and the outcome (up: increase in 70% or more of relevant results; down: decrease in 70% or more of relevant results; horizontal: study provides inconsistent evidence of effect direction). The shading of the arrow denotes whether the association was statistically significant (filled: p≤.05 in 70% or more of relevant results; unfilled: p≥.05 in 70% or more of relevant results; grey: study provides inconsistent evidence of statistical significance).

**Supporting information S4.** Descriptive characteristics of included studies conducted within a retail setting, by outcome, quality and presence of confounding promotions

| **Outcome category** | **Author, publication year, country** | **Study design** | **Sample size** | **Promotions used (targeted products)** | **Confounding promotions** | **Direction of effect ^a^** | **Study quality** |
| --- | --- | --- | --- | --- | --- | --- | --- |
| Sales (electronically recorded) | Daunfeldt, 2014, Sweden^70^ | Observational (11 months pre-, 7 months post implementation) | 1 supermarket (~13,338 transactions) | Shelf labels (organic olive oil, coffee and flour) | None | ◄► | Strong |
| Sales (‘take-home’ panel) | Nakamura, 2014, UK^73^ | Observational (cross-sectional, 12 months of aggregated sales data) | 1 supermarket (5,787 transactions) | End-of-aisle displays (carbonated drinks, coffee and tea) | None | ▲ | Strong |
| Sales (electronically recorded) | Harris, 2020, USA^90^ | Observational (cross-sectional, 12 months aggregated sales data) | 2,321 products, supermarkets in 8 cities | End-of-aisle, front-of-store and in-aisle displays (fruit flavoured soft drinks) | None | ▲ | Moderate |
| Sales (electronically recorded) | Payne, 2018, USA^68^ | Non-randomised, controlled study (1 month pre-, 1 month post implementation) | 3 supermarkets (1 control, 2 intervention) | Checkout placement and verbal prompts (fruit and vegetables) | None | ▲ | Moderate |
| Sales (electronically recorded) | Van Gestel, 2018, Netherlands^79^ | Uncontrolled pre-post (1 month pre-, 1 month post implementation) | 1 kiosk | Checkout placement (fruit, nuts, muesli bars, cereal biscuits, crackers) | None | ▲ | Moderate |
| Sales (sales receipts) | Gustafson, 2019, USA^60^ | Non-randomised, controlled study (repeated cross-section baseline and post implementation) | 497 individuals, 16 supermarkets (6 control, 10 intervention) | End-of-aisle placement, recipe cards, grocery cart placards, branded banners (healthy recipes including fruit and vegetables) | Taste testing, price reductions | △ | Moderate |
| Sales (electronically recorded) | Hua, 2017, USA^39^ | Randomised controlled study (12 month baseline, 5 month intervention period) | 14 vending machines (8 control, 6 intervention) | Vending machine promotional stickers (healthy snacks and beverages) | Nutritional labelling | ◁▷^b^ | Moderate |
| Sales (electronically recorded) | Toft, 2017, Denmark^88^ | Non-randomised, controlled study (1 month baseline, 3 month intervention period) | 4 supermarkets (3 control, 1 intervention) | Entrance displays, checkout display, island display, End-of-aisle displays (fruit and vegetables) | Availability | ◁▷ | Moderate |
| Sales (researcher recorded) | Caspi, 2017, USA^76^ | Observational (cross-sectional) | 594 individuals, 99 small food stores | Entrance displays, checkout displays (fruit, vegetables and whole grains) | None | △ | Weak |
| Sales (electronically recorded) | Garrido-Morgado 2015, Spain^84^ | Observational (cross-sectional analysis, 10 weeks of aggregated sales data) | 1 supermarket | End-of-aisle displays, island displays (milk) | None | ▲ | Weak |
| Sales (participant report) | Salmon, 2015, Netherlands^55^ | Randomised controlled study (cross-sectional between-subjects) | 127 individuals, 1 supermarket | In-premise posters (low-fat cheese) | None | △ | Weak |
| Sales (participant report) | Sanchez-Flack, 2017, USA^45^ | Observational (cross-sectional) | 369 individuals, 16 convenience stores | End-of-aisle displays, checkout displays, Entrance displays (fruit and vegetables) | None | ▲ | Weak |
| Sales (electronically recorded) | Sigurdsson, 2014, Norway^54^ | Uncontrolled pre-post (3-4 weeks pre-, 9-10 weeks post implementation) | 1 convenience store, 1 supermarket (102,368 transactions) | Checkout displays, shelf labels (dried fish, dried fruit and nut mix) | None | △^b^ | Weak |
| Sales (electronically recorded) | Winkler, 2016, Denmark^71^ | Non-randomised, controlled study (4 weeks pre, 4 weeks post implementation) | 18 supermarkets (14 control, 4 intervention) | Checkout displays (fresh fruit, fruit bars, carrots, dried fruit) | None | △ | Weak |
| Sales (storeowner report) | Budd, 2017, USA^58^ | Randomised controlled study (retrospective estimate of sales data at baseline and post implementation) | 12 convenience stores (6 control, 6 intervention) | In-premise posters, branded refrigerators, shelf labels, bags (water, low calorie soft drinks, milk, bread, fish, fruit, vegetables, low fat snacks) | Taste testing, availability | ▽ | Weak |
| Sales (electronically recorded) | Chapman, 2019, USA^64^ | Non-randomised, controlled study (14 weeks pre, 4 weeks post implementation) | 4 stores (2 control, 2 intervention) | In-premise posters, shelf labels, floor arrows (fruit, granola bars) | Placement within aisle | ◁▷ | Weak |
| Sales (participant report) | Dannefer, 2012, USA^33^ | Uncontrolled pre-post (repeated cross-section at baseline and post implementation) | 617 individuals, 8 convenience stores | In-premise posters, branded refrigerators (water, low fat milk, low-sodium canned goods, whole grain bread, healthy snacks and sandwiches) | Cooking demonstrations, recipe giveaways, availability, placement within shelf | △^b^ | Weak |
| Sales (electronically recorded) | Fiske, 2004, USA^38^ | Randomised controlled study (2 weeks pre, 4 weeks post implementation) | 10 vending machines (2 control, 8 intervention) | In-premise posters, shelf labels (low fat snacks) | Placement within shelf, availability | △ | Weak |
| Sales (electronically recorded) | Foster, 2014, USA^36^ | Randomised controlled study (3 months pre, 6 months post implementation) | 4 intervention, 4 control supermarkets | In-premise posters, shelf labels (milk, cereal, frozen meals, low calorie soft drinks and water) | Placement within shelf, availability, co-location of complementary products, taste testing, free samples | ▲ | Weak |
| Sales (manually recorded) | French, 2001, USA^34^ | Randomised controlled study (cross-sectional analysis, 1 month aggregated sales data) | 24 school/workplace sites (8 control, 12 intervention), 55 vending machines | Vending machine signage promoting value (low fat snacks) | Availability | ▲ | Weak |
| Sales (electronically recorded) | Gamburzew, 2016, France^78^ | Non-randomised, controlled study (within-subject pre- and post-implementation) | 6,625 individuals, 4 supermarkets (2 control, 2 intervention) | Shelf labels, in-premise posters, leaflets, prime placement (foods of good nutritional value according to SAIN/LIM model) | Taste testing | △ | Weak |
| Sales (participant report) | Kristal, 1997, USA^37^ | Randomised controlled study (repeated cross-section at baseline and post implementation) | 960 individuals, 8 supermarkets (4 control, 4 intervention) | In-premise posters, leaflets (fruit and vegetables) | Money-off coupons, food demonstrations | △ | Weak |
| Sales (researcher recorded) | Lawman, 2015, USA^66^ | Observational (repeated cross-section, baseline and post implementation) | 14,620 individuals, 113 convenience stores | In-premise posters, shelf labels, recipe cards (fruit and vegetables, low fat dairy, meat, whole grains) | Availability | ◁▷ | Weak |
| Sales (researcher recorded) | Paine-Andrews, 1996, USA^89^ | Uncontrolled pre-post (multiple baseline and intervention periods) | 1 supermarket | Verbal prompts (low fat milk, salad dressing, frozen desserts) | Taste testing, price | △^b^ | Weak |
| Sales (electronically recorded) | Pharis, 2018, USA^48^ | Uncontrolled pre-post (12 months pre, 12 months post implementation) | ~250 vending machines | Vending machine signage (low calorie drinks, fruit juice, low fat snacks) | Availability, price, position within shelf, nutritional labelling | ▲ | Weak |
| Sales (electronically recorded) | Sigurdsson, 2011, Norway^85^ | Uncontrolled pre-post (multiple baseline and intervention periods) | 2 stores | Checkout placement, in-premise posters (bananas) | Position within shelf | △ | Weak |
| Sales (participant report) | Trude, 2018, USA^43^ | Randomised controlled study (within-subjects pre-post implementation) | 509 individuals, 30 communities (16 control, 14 intervention) | Checkout placement, in-premise posters, leaflets (low and sugar and/or high fibre snacks and beverages) | Availability, community education sessions, taste testing, social media campaign | ▲ | Weak |
| Consumption (participant report) | Ayala, 2013, USA^40^ | Randomised controlled study (within-subjects pre-post implementation) | 119 individuals, 4 convenience stores (2 control, 2 intervention) | Point-of-sale promotion, in-premise posters, shelf labels, streamers, checkout display, audible promotion (fruit and vegetables) | Nutritional labelling, taste testing | △ | Weak |
| Consumption (participant report) | Kristal, 1997, USA^37^ | Randomised controlled study (repeated cross-section at baseline and post implementation) | 960 individuals, 8 supermarkets (4 control, 4 intervention) | In-premise posters, leaflets (fruit and vegetables) | Money-off coupons, food demonstrations | △ | Weak |
| Consumption (participant report) | Trude, 2018, USA^43^ | Randomised controlled study (within-subjects pre-post implementation) | 509 individuals, 30 communities (16 control, 14 intervention) | Checkout placement, in-premise posters, leaflets (low and sugar and/or high fibre snacks and beverages) | Availability, community education sessions, taste testing, social media campaign | ◁▷ | Weak |
| Customer attention (eye tracking) | Hurley, 2017, USA^57^ | Non-randomised, controlled study (cross-sectional between-subjects) | 89 individuals | Shelf labels (carbonated soft drink) | None | ▲ | Weak |
| Health outcomes (BMI) | Cohen, 2015, USA^59^ | Observational (cross-sectional) | 980 individuals, 40 food stores | End-of-aisle displays, special floor displays, checkout displays (sugar sweetened beverages, foods high in solid fats, fruit, vegetables and whole grain products) | None | ▲ | Moderate |
| Diet quality (participant report) | Cohen, 2015, USA^59^ | Observational (cross-sectional) | 980 individuals, 40 food stores | End-of-aisle displays, special floor displays, checkout displays (sugar sweetened beverages, foods high in solid fats, fruit, vegetables and whole grain products) | None | ◁▷^c^ | Moderate |
| Diet quality (participant report) | Baird, 2018, UK^44^ | Non-randomised, controlled study (between-subjects pre-post implementation) | 138 individuals | Entrance displays, checkout displays (fruit and vegetables) | Placement within supermarket | ▲ | Weak |
| Diet quality (participant report) | Trude, 2018, USA^43^ | Randomised controlled study (within-subjects pre-post implementation) | 509 individuals, 30 communities (16 control, 14 intervention) | Checkout placement, in-premise posters, leaflets (low and sugar and/or high fibre snacks and beverages) | Availability, community education sessions, taste testing, social media campaign | ◁▷ | Weak |

^a^ Orientation of arrow denotes the direction of association between the promotion and the outcome (up: increase in 70% or more of relevant results; down: decrease in 70% or more of relevant results; horizontal: study provides inconsistent evidence of effect direction). The shading of the arrow denotes whether the association was statistically significant (filled: p≤.05 in 70% or more of relevant results; unfilled: p≥.05 in 70% or more of relevant results; grey: study provides inconsistent evidence of statistical significance).

^b^ No null-hypothesis significance testing was conducted for the comparison of interest.

^c^ Direction of effect not reported.

**Supporting information S5.** Descriptive characteristics of included studies conducted within an out-of-home setting, by outcome, quality and presence of confounding promotions

| **Outcome category** | **Author, publication year, country** | **Study design** | **Sample size** | **Promotions used (targeted products)** | **Confounding promotions** | **Direction of effect ^a^** | **Study quality** |
| --- | --- | --- | --- | --- | --- | --- | --- |
| Sales (electronically recorded) | van Kleef, 2015, Netherlands^67^ | Uncontrolled pre-post (10 weeks pre- 1-8 weeks post implementation) | 1 restaurant (11,519 transactions) | Verbal prompts (Pancakes, orange juice, fruit salad) | None | ▲ | Strong |
| Sales (recorded by observer) | Saulais, 2019, France^53^ | Uncontrolled pre-post (multiple baseline and intervention periods) | 1 restaurant, 294 individuals | Verbal prompt, text on menu (vegetarian burger, meatballs) | None | ▲ | Moderate |
| Sales (electronically recorded) | Wagner, 1988, USA^42^ | Uncontrolled pre-post (multiple baseline and intervention periods) | 2 restaurants (1 control, 1 intervention) | In-premise posters, table tents (Salad) | None | △^b^ | Moderate |
| Sales (sales receipts) | Lee-Kwan, 2015, USA^52^ | Randomised controlled study (1 month baseline, 7 month intervention period) | 7 carry out restaurants, 4 control, 3 intervention (186,654 transactions) | Text and images on menus, in-premise posters (47 low calorie, low fat menu items) | Availability | ▲ | Moderate |
| Sales (electronically recorded) | Anzman-Frasca, 2018, USA^32^ | Randomised controlled study (multiple baseline and intervention periods) | 1 restaurant, 58 individuals | Verbal prompts, images on menus (healthy child meals) | None | △ | Weak |
| Sales (electronically recorded) | Broers, 2019, Belgium^61^ | Non-randomised, controlled study (multiple baseline and intervention periods) | 2 restaurants (~1860 transactions) | In-premise posters (vegetable soup) | None | ▲ | Weak |
| Sales (electronically and manually recorded) | Buscher, 2001, Canada^86^ | Uncontrolled pre-post (multiple baseline and intervention periods) | 1 cafeteria (~277,728 transactions) | In-premise posters (fruit, vegetables, pretzels, yoghurt) | None | ▲ | Weak |
| Sales (electronically recorded) | Chapman, 2012, UK^69^ | Uncontrolled pre-post (multiple baselines, one intervention period) | 1 canteen (1,598 transactions) | Checkout placement (confectionery, biscuits, fruit) | None | ▼ | Weak |
| Sales (electronically recorded) | Cheung, 2019, Netherlands^91^ | Uncontrolled pre-post (1 week baseline, 2 weeks intervention) | 1 canteen (51,198 transactions) | Checkout placement, in-premise posters, presentation bin (fruit, bread) | None | ▲ | Weak |
| Sales (recorded by observer) | Collins, 2019, UK^49^ | Uncontrolled pre-post (1 week baseline, 2 weeks intervention) | 2 canteens (11,650 transactions) | In-premise posters (vegetables) | None | ▲ | Weak |
| Sales (recorded by observer) | Ebster, 2006, Austria^80^ | Non-randomised, controlled study (post implementation only) | 1 restaurant (2,160 transactions) | Verbal prompts (potato salad, fries) | None | ▲ | Weak |
| Sales (electronically recorded) | Ayala, 2017, USA^30^ | Randomised controlled study (2 months post implementation only) | 8 restaurants (4 control, 4 intervention) | Table tents, in-premise posters, menu design (healthy child meals) | Out-of-scope placement | △ | Weak |
| Sales (electronically recorded) | Ensaff, 2015, UK^87^ | Non-randomised, controlled study (9.5 months pre-, 9.5 months post-implementation) | 2 school canteens (1 control, 1 intervention) | Checkout placement, in-premise posters, shelf talkers (vegetarian menu items, salad sandwiches, fruit) | Packaging | ▲ | Weak |
| Sales (electronically recorded) | Fitzgerald, 2004, USA^83^ | Uncontrolled pre-post (1 month baseline, 1 month post implementation) | 9 restaurants (53,692 transactions) | In-premise posters, table tents (menu items meeting nutritional guidelines) | Out-of-premise marketing campaign | △ | Weak |
| Sales (electronically recorded) | Lopez, 2017, USA^65^ | Uncontrolled pre-post (1 month baseline, 2 months post implementation) | 4 restaurants (10,664 transactions) | Verbal prompts, in-premise posters, placemats (menu items meeting nutritional guidelines) | Free gift (toy), out-of-premise marketing campaign | ◄► | Weak |
| Sales (recorded by observer) | Mistura, 2019, Canada^63^ | Uncontrolled pre-post (multiple baseline and intervention periods) | 1 canteen (24,410 transactions) | In-premise posters (vegetables) | Position on buffet line, nutrition information | △ | Weak |
| Sales (participant report) | Trude, 2018, USA^43^ | Randomised controlled study (within-subjects pre-post implementation) | 509 individuals, 30 communities (16 control, 14 intervention) | Checkout placement, in-premise posters, leaflets (low and sugar and/or high fibre snacks and beverages) | Availability, community education sessions, taste testing, social media campaign | ▲ | Weak |
| Sales (participant report) | Wolfenden, 2015, Australia^72^ | Randomised controlled study (within-subjects pre-post implementation) | 85 sports clubs (43 control, 42 intervention), 1,143 individuals | In-premise posters (fruit and vegetables, low calorie beverages) | Availability, meal deal, price | ▲ | Weak |
| Consumption (plate waste measurement) | Anzman-Frasca, 2018, USA^32^ | Randomised controlled study (multiple baseline and intervention periods) | 1 restaurant, 57 individuals | Verbal prompts, images on menus (healthy child meals) | None | ◁▷^c^ | Weak |
| Consumption (observer recorded) | Schwartz, 2007, USA^51^ | Non-randomised, controlled study (cross sectional, post implementation only) | 2 schools (1 control, 1 intervention), ~323 individuals | Verbal prompt (fruit) | None | △ ^b^ | Weak |
| Consumption (participant report) | Trude, 2018, USA^43^ | Randomised controlled study (within-subjects pre-post implementation) | 509 individuals, 30 communities (16 control, 14 intervention) | Checkout placement, in-premise posters, leaflets (low and sugar and/or high fibre snacks and beverages) | Availability, community education sessions, taste testing, social media campaign | ◁▷ | Weak |
| Consumer preference (menu items chosen) | Dos Santos, 2020, Denmark, France, Italy, UK^47^ | Non-randomised, controlled study (cross sectional, post implementation only) | 360 individuals | Verbal prompt, Text on menu (vegetarian meatballs) | None | △ | Moderate |
| Consumer preference (menu items chosen) | Bacon, 2018, UK^41^ | Randomised controlled study (cross-sectional between-subjects) | 750 individuals | Text on menu, menu design (risotto, ravioli) | None | ◁▷ | Weak |
| Consumer preference (participant-reported purchase intentions) | Gala, 2018, USA^31^ | Randomised, controlled study (cross-sectional between-subjects) | 416 individuals | Images on menu (Burger, salad) | None | ▼ | Weak |
| Consumer preference (participant-reported purchase intentions) | Hou, 2017, USA^81^ | Randomised, controlled study (cross-sectional between-subjects) | 315 individuals | Images on menu (chocolate ice-cream, chicken and egg salad) | None | ▲ | Weak |
| Consumer preference (participant-reported purchase intentions) | Nazlan, 2018, USA^56^ | Randomised, controlled study (cross-sectional between-subjects) | 592 individuals | Text on menu (chicken sandwich) | None | ◁▷ | Weak |
| Consumer preference (observer recorded) | Schwartz, 2007, USA^51^ | Non-randomised, controlled study (cross sectional, post implementation only) | 2 schools (1 control, 1 intervention), ~323 individuals | Verbal prompt (fruit) | None | △^b^ | Weak |
| Diet quality (participant report) | Trude, 2018, USA^43^ | Randomised controlled study (within-subjects pre-post implementation) | 509 individuals, 30 communities (16 control, 14 intervention) | Checkout placement, in-premise posters, leaflets (low and sugar and/or high fibre snacks and beverages) | Availability, community education sessions, taste testing, social media campaign | ◁▷ | Weak |

^a^ Orientation of arrow denotes the direction of association between the promotion and the outcome (up: increase in 70% or more of relevant results; down: decrease in 70% or more of relevant results; horizontal: study provides inconsistent evidence of effect direction). The shading of the arrow denotes whether the association was statistically significant (filled: p≤.05 in 70% or more of relevant results; unfilled: p≥.05 in 70% or more of relevant results; grey: study provides inconsistent evidence of statistical significance).

^b^ No null-hypothesis significance testing was conducted for the comparison of interest.

^c^ Direction of effect not reported.

**Supporting information 6.** Descriptive characteristics of included studies conducted within an online shopping environment, by outcome, quality and presence of confounding promotions

| **Outcome category** | **Author, publication year, country** | **Study design** | **Sample size** | **Promotions used (targeted products)** | **Confounding promotions** | **Direction of effect ^a^** | **Study quality** |
| --- | --- | --- | --- | --- | --- | --- | --- |
| Sales (electronically recorded) | Delaney, 2017, Australia^35^ | Randomised, controlled study (within-subjects pre-post implementation) | 10 schools (5 control, 5 intervention) 2,714 individuals | Position on front page, prompts (lower energy, saturated fat, sugar and salt alternatives) | Availability, nutritional labelling, position within menu | ◄► | Moderate |
| Sales (electronically recorded) | Hou, 2017, USA^82^ | Randomised, controlled study (cross-sectional between-subjects) | 91 individuals | Text prompt (cookies) | None | ▼ | Weak |
| Consumer preference (menu items chosen) | Forwood, 2015, UK^75^ | Randomised, controlled study (cross-sectional between-subjects) | 720 individuals | Pop-up prompt (lower energy density alternatives) | None | △ | Moderate |
| Consumer preference (menu items chosen) | Koutoukidis, 2018, UK^62^ | Randomised, controlled study (cross-sectional between-subjects) | 1,088 individuals | Pop-up prompt (lower saturated fat alternatives) | None | ▲ | Moderate |
| Consumer preference (menu items chosen) | Breugelmans, 2005, Netherlands^50^ | Non-randomised, controlled study (cross-sectional between-subjects) | 387 individuals | Position on front page (margarine, cereal) | None | ◄► | Weak |
| Consumer preference (menu items chosen) | Nederkoorn, 2014, Netherlands^77^ | Randomised, controlled study (cross-sectional between-subjects) | 118 individuals | On-screen advertisements (pizza, crisps, cookies, candy) | None | ◁▷^b^ | Weak |

^a^ Orientation of arrow denotes the direction of association between the promotion and the outcome (up: increase in 70% or more of relevant results; down: decrease in 70% or more of relevant results; horizontal: study provides inconsistent evidence of effect direction). The shading of the arrow denotes whether the association was statistically significant (filled: p≤.05 in 70% or more of relevant results; unfilled: p≥.05 in 70% or more of relevant results; grey: study provides inconsistent evidence of statistical significance).

^b^ Direction of effect not reported.
