## Supplementary material for "The Impact of Non-Price In-premise Marketing on Food and Beverage Purchasing and Consumer Behaviour: A Systematic Review": Table 1. Inclusion and exclusion criteria

| **Table 1.** Inclusion and exclusion criteria | |
| --- | --- |
| **Inclusion criteria** | **Exclusion criteria** |
| Examines exposure or intervention altering in-premise positioning or advertising of food or drink in retail, out-of-home or online setting | Qualitative methodology |
| Recorded outcomes relating to sales, consumer preference, attention, dietary quality or health status | Lack of primary data |
| Conducted in a high-income country according to world-bank criteria | Existing systematic or narrative review |
| Published in English | Focus on out-of-premise promotion (e.g., television, billboards or printed media) |
