## Supplementary material for "The Impact of Non-Price In-premise Marketing on Food and Beverage Purchasing and Consumer Behaviour: A Systematic Review": Figure 1. PRISMA flow diagram

**Screening**

**Included**

**Eligibility**

**Identification**

Records identified through database searching
n = 21,679

Additional records identified through other sources
n = 1

Records after duplicates removed
n = 19,285

Full-text articles assessed for eligibility
n = 190

Studies included in vote-counting synthesis
n = 62

Title and abstract screening
n = 19,285

Records excluded
n = 19,095

Full-text articles excluded

(n=128)
n = 8 unobtainable

n = 8 duplicate

n = 1 non-food product

n = 9 wrong type of study

n = 32 wrong/no outcome

n = 9 wrong setting

n = 61 wrong/no promotion

**Figure 1.** PRISMA flow diagram.
